# Socio-economic Disparities in Adult Mortality in Latin America*

**DOI:** 10.1101/2024.11.23.24317823

**Authors:** Giancarlo Buitrago, Sofía Marinkovic Dal Poggetto, Antonella Bancalari, Samuel Berlinski, Dolores de la Mata, Marcos Vera-Hernández

## Abstract

**Introduction:** Large socioeconomic disparities in health care utilization and health outcomes have been well-documented in Latin American countries. However, little is known about disparities in mortality rates. We estimate socioeconomic gradients in mortality in the Latin American region and discuss their patterns.

**Methods:** We utilize death certificate data from the national vital statistics systems and population data from national censuses in Argentina, Brazil, Chile, Colombia, Ecuador, Mexico, and Peru (2010-2019) to calculate mortality rates by age, sex, and educational attainment. We also calculate mortality rates by cause of death. Data are harmonized to ensure comparability across countries and between death certificates and census data within countries. To analyze socioeconomic disparities, we compute the ratio between the mortality rate for individuals with a lower level of education (secondary incomplete or less) and the mortality rate for individuals with a higher level of education (secondary complete or more) by age and sex. The socioeconomic analysis is limited to adults aged 20 years or older.

**Results:** Mortality rates for individuals with lower education are higher at all age groups than for individuals with higher education, with larger disparities observed in younger age groups. Differences across countries in these inequalities are also more pronounced in younger cohorts. In the 20-69 age range, highly educated groups show similar mortality rates across countries, with most differences occurring in lower education groups. Lower education is associated with higher mortality rates from violent causes, particularly before age 50. Among non-violent causes, infectious diseases exhibit larger socioeconomic gradients than non-communicable diseases. Amongst the latter, deaths from diabetes and cardiovascular diseases exhibit more socioeconomic inequality than those from neoplasms.

**Conclusion:** Despite overall improvements in average health indicators in the region, which are concomitant to a fall in income inequality and expansion of universal health coverage, significant challenges remain in addressing disparities in mortality rates, particularly for younger populations and women.

**Key Messages:**

- **What is already known on this topic**

Latin America has long been recognized for substantial socioeconomic disparities in key health outcomes, including life expectancy and infant mortality. While previous studies have focused primarily on specific age groups (e.g., infants or the elderly) or aggregated levels (e.g., countries or select cities), there is limited evidence on socioeconomic inequalities in overall mortality rates across the life course or for different causes of death, mainly using individual-level data.

- **What this study adds**

This study comprehensively analyses socioeconomic disparities in mortality across seven Latin American countries between 2010 and 2019. Using detailed microdata from censuses and death certificates, it highlights mortality disparities by educational attainment, age, and sex, revealing a clear educational gradient that diminishes with advancing age. These findings offer novel insights into health inequities in the region.

- **How this study might affect research, practice or policy**

Our findings underscore the need for targeted interventions and policies to address persistent mortality inequalities in Latin America, particularly among the most vulnerable populations. The observed patterns also provide a basis for future research to explore the drivers of these disparities and evaluate the impact of social and health policies on reducing inequalities in the region.

## 1 INTRODUCTION

Latin America remains one of the most unequal regions globally, where deep-rooted socioeconomic disparities significantly impact key health outcomes. These inequalities are consistently observed across various dimensions, including life expectancy, infant mortality, and cause-specific mortality rates.^1, 2, 3, 4, 5^ Despite considerable progress in expanding healthcare coverage and access, these disparities persist, possibly driven by structural inequalities ingrained in the region’s social and economic systems.^6, 7^ These inequalities are evident between countries and within them, with stark differences observed across urban and rural areas and between different socioeconomic groups. This persistent inequality underscores the critical need for ongoing research and targeted policy interventions to address Latin America’s underlying social determinants of health.

Most existing studies on mortality inequalities in Latin America have focused on single countries, primarily investigating infant and maternal mortality, often relying on surveys such as the Demographic and Health Surveys (DHS) and UNICEF Multiple Indicator Cluster Surveys (MICS).^8, 9, 2, 10^ While these studies have been crucial in identifying specific disparities, they provide a limited understanding of the broader mortality patterns across the life course and various health outcomes. The SALURBAL research group has offered valuable insights into urban health disparities in Latin American cities in recent years.^1, 11, 12^ However, over the past decade, there has been a notable gap in research that systematically and comprehensively compares adult mortality inequalities within and across multiple Latin American countries, particularly concerning all-cause mortality or disease-specific causes and how they vary at different ages. Addressing this gap is essential for a complete understanding of health disparities in the region and for informing targeted public health interventions.

This study aims to provide a comprehensive and systematic analysis of socioeconomic inequalities in adult mortality across 7 Latin American countries: Argentina, Brazil, Chile, Colombia, Ecuador, Mexico, and Peru. By focusing on individuals aged 20 and above, we ensure that the population studied has a higher likelihood of completing or being near the completion of their formal education. This approach allows for a more robust link between individual educational attainment and age-adjusted mortality rates. A novel aspect of this research is using the most recent census data, combined with death certificates from the year immediately following the census, enabling precise, fine-grained within and cross-country comparisons. Our findings offer critical insights into persistent health disparities across various causes of death, age groups, and sexes, demonstrating that these inequalities persist despite significant socioeconomic changes and health system reforms in the region. These results have the potential to significantly influence targeted public health policies and interventions aimed at reducing health disparities and promoting equity in health outcomes throughout Latin America.

## 2 METHODS

### 2.1 Study Design and Population

We study socioeconomic disparities in mortality in 7 Latin American countries: Argentina, Brazil, Chile, Colombia, Ecuador, Mexico, and Peru. We selected these countries based on the availability of educational attainment information in the death certificates and population census.

Mortality rates for any given group (i.e., education-age-sex combination) are calculated as the ratio between the number of deaths in period t+1 of that group and the population count of that same group in period t, as this corresponds to the population at risk.

We follow the literature in using educational attainment as a measure of socio-economic status.^13, 14, 15, 16, 17^ We divide the population into two groups: i) those with completed primary education (which may include people with secondary education incomplete) or less, and ii) those with completed secondary education or more. Because we use education as a measure of socio-economic status, we focus on the population 20 and older when secondary education must have been completed.

We calculate mortality rates by age group, sex, and educational attainment. We begin by analyzing disparities in all causes of death. We then examine inequalities by cause of death: communicable, maternal and nutritional, vs. non-communicable diseases.^18^ Finally, we compare socio-economic gradients between intentional (i.e., violent) versus unintentional injuries.

Informed consent was not required because we used anonymized and retrospective data.

### 2.2 Data

We use death certificate data from the national vital statistics systems and population data from censuses in our analysis (see Appendix, Table S1, for details on the sources). The year of analysis for each country is determined by the most recent population census available: Argentina (2011), Brazil (2011), Chile (2018), Colombia (2019), Ecuador (2011), Mexico (2011), and Peru (2018).

For 6 of the 7 countries in our study we obtain population counts by age, sex and educational attainment from the entire population censuses. The exception is Brazil, where the educational attainment variable is only available for a sub-sample of residents. We use IPUMS-International 10% public-use microdata to estimate population counts for this country.

We utilize the international classification codes (ICD-10) to identify the cause of death. We considered various causes of death –either as the primary or secondary cause. We follow closely the WHO classification of ICD-10 codes into the following disease groups:^18^ (i) communicable, maternal, nutritional, (ii) non-communicable, (iii) intentional and (iv) unintentional injuries. Within non-communicable mortality, we further focus on cardiovascular diseases, diabetes and neoplasms, as these conditions have increasingly ranked among the leading causes of disease burden in the region.^19, 5^ We also categorize intentional injuries into homicides and suicides to differentiate between crime-related and mental health-related root causes.

Data are harmonized to ensure comparability across countries and between death certificates and census data within countries (see Appendix for details). Education categories were defined to construct analogous categories across each country and database. The two categories, which refer to the maximum educational attainment, are: i) Primary education complete (which may include individuals with incomplete secondary education) or less, and ii) Secondary education complete or more. We refer to these two levels as low and high education levels, respectively. Education categories from death certificate registries and census data were harmonized to align with these two education levels (see Table S2 in the Appendix for a detailed description of the variable construction).

The quality of death certificate registries in the countries analyzed has been classified as medium to high, with the exception of Peru, which is classified as low-quality.^201^ Estimates from the Pan American Health Organization^21^ indicate that the under-reporting of deaths in the years considered in this study amounts to 20% in Ecuador (2011), and 29% in Peru (2018). Under-reporting estimates are much lower in other countries: around 13% for Colombia (2019) and below 10% in the rest of the countries, with Argentina (2011) showing no under-reporting issues. Appendix subsection 7.1.5 reports the percentage of death certificates which have missing information on education and cause of death.

### 2.3 Statistical Analysis

We study inequality in mortality by constructing mortality rates based on age and sex and across low and high educational attainment (as defined above). Additionally, we examine these differences by cause of death.

Mortality rates are expressed as deaths per 1,000 population to be comparable with national statistics. The rates are obtained as a simple ratio between death and population counts:

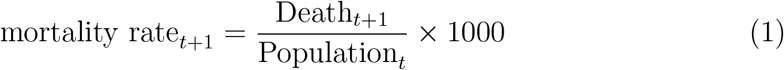

We measure socioeconomic inequality by age and sex as the ratio of the mortality rate of the least educated to the most educated:

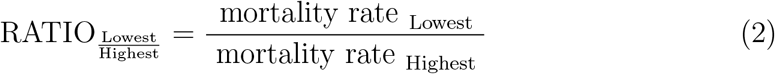

A ratio greater than one means that the mortality rate of the less educated is higher than that of the more educated. We compute these measures for all deaths and by specific cause of death.

We assume that the distribution of educational attainment among individuals with missing educational attainment in the death certificate is the same as the distribution observed in the census for individuals of the same age and sex. For Peru, 28% of the death certificates have cause of death missing and were excluded from the cause of death analysis. Appendix subsection 7.1.5 provides more detailed information on missing information.

### 2.4 Funding

This paper has been partially funded by the Latin America and Caribbean Inequality Review (https://lacir.lse.ac.uk/).

## 3 RESULTS

### 3.1 General Results

Our study covers seven Latin American countries over ten years. These countries are characterized by relatively high life expectancy at birth rates (see Table 1). For females (males), the rate in 2019 ranged from 77.6 (70.9) in Mexico to 82.7 (77.9) in Chile. The differences in longevity between men and women translate into a significantly larger share of the population over 65 that is female in the countries we study. The analysis of mortality rates that follows should be assessed against the backdrop of these statistics.

**Table 1:** Life expectancy at birth & share of population 65+ years old (UN)

| Country | Year | Life Expectancy at birth |  | Population +20 (%) |  | Population +65 (%) |  |
| --- | --- | --- | --- | --- | --- | --- | --- |
|  |  | Female | Male | Female | Male | Female | Male |
| Argentina | 2011 | 79.3 | 72.8 | 67.3 | 64.8 | 12.5 | 8.7 |
| Argentina | 2019 | 80.7 | 73.9 | 69.4 | 67.1 | 13.6 | 9.6 |
| Brazil | 2011 | 76.7 | 70.1 | 68.1 | 66.0 | 7.9 | 6.2 |
| Brazil | 2019 | 78.5 | 72.2 | 72.2 | 70.1 | 10.1 | 7.9 |
| Chile | 2011 | 81.7 | 76.5 | 71.5 | 69.9 | 11.2 | 8.7 |
| Chile | 2019 | 82.7 | 77.9 | 75.2 | 73.9 | 13.4 | 10.9 |
| Colombia | 2011 | 78.5 | 72.1 | 65.0 | 62.9 | 6.8 | 5.5 |
| Colombia | 2019 | 79.7 | 73.8 | 70.3 | 68.3 | 9.0 | 7.4 |
| Ecuador | 2011 | 78.7 | 72.9 | 60.2 | 58.7 | 6.6 | 5.4 |
| Ecuador | 2019 | 80.0 | 74.7 | 64.8 | 63.3 | 8.0 | 6.8 |
| Mexico | 2011 | 77.3 | 71.5 | 62.2 | 59.6 | 6.6 | 5.9 |
| Mexico | 2019 | 77.6 | 70.9 | 66.7 | 64.3 | 8.3 | 7.4 |
| Peru | 2011 | 76.3 | 72.1 | 61.7 | 60.8 | 7.6 | 7.1 |
| Peru | 2019 | 78.5 | 73.9 | 65.3 | 63.6 | 8.6 | 7.7 |

*Note:* Data comes from United Nations, Department of Economic and Social Affairs, Population Division (2024). World Population Prospects 2024, Online Edition.

### 3.2 Socioeconomic Disparities in Mortality Rates

Ratios of adult mortality rates between low and high education are reported by sex and age group in Figure 1, and Appendix Tables S7 and S8. Inequalities in mortality rates are pronounced across sex and age. Inequalities are larger in younger age groups than in older ones. For women (men) the unweighted median inequality ratio by age group for our sample of countries falls continuously from 1.93 (2.27) in the 20-29 age group to 1.39 (1.22) in the 80+ age group.

**Figure 1.**
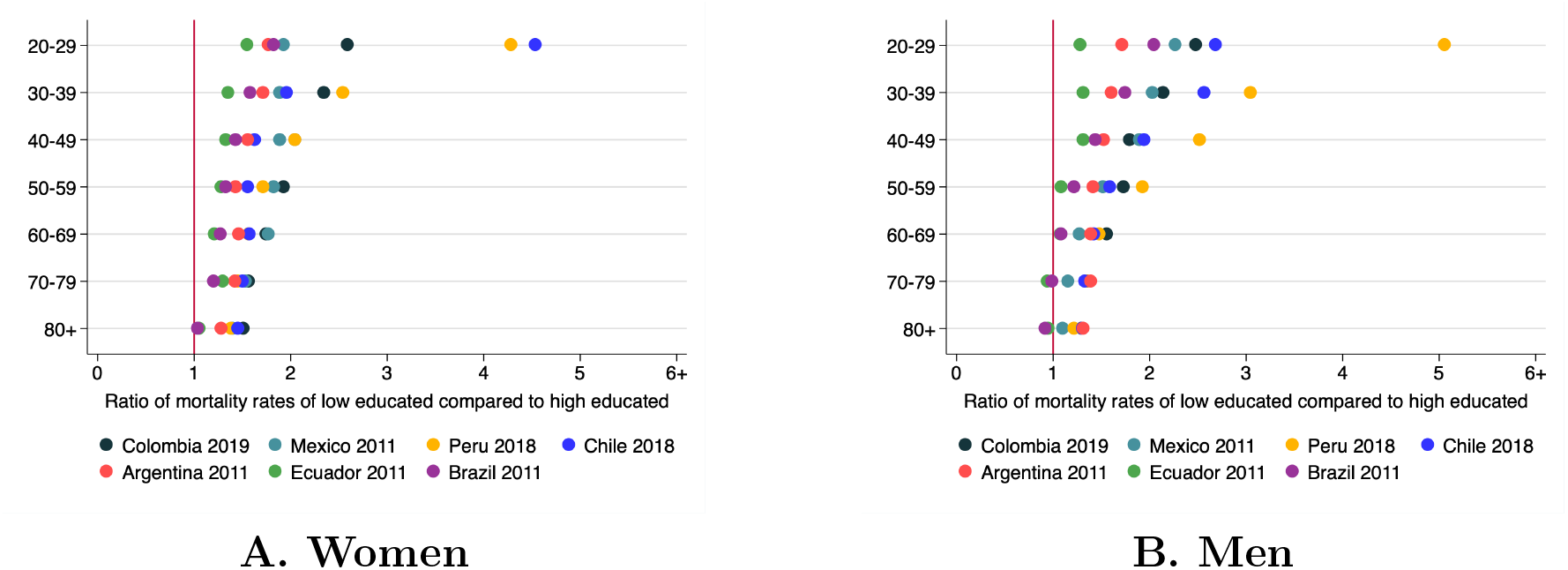
Socioeconomic inequality in mortality, by age and sex

The pattern of socioeconomic inequality in mortality also differs by sex. At younger ages, the unweighted median mortality ratio is larger for men than women, but the opposite is true for the 50-59 age group and older ones.

The variability of socio-economic mortality ratios across countries is much larger in younger than older age groups. It is evident that the greatest disparities in the 20-29 age group are found in Peru (2018) and Chile (2018) for both women and men, where the mortality rates of the least educated are more than three times higher than those for the most educated and can be up to 5 times larger (men in Peru). In contrast, the largest mortality ratio for the 80+ age group is 1.54 (women in Chile).

#### 3.2.1 Socioeconomic Disparities in Mortality Rates in Communicable, Maternal and Nutritional versus Non-communicable Diseases

Communicable, maternal, nutritional and non-communicable diseases represent 86.23 percent of all the deaths in our sample (see Appendix Table S6). Across all countries in our analysis, the majority of mortality is due to non-communicable diseases (a percentage as large as 83.3% in Chile in 2018) rather than communicable, maternal, and nutritional diseases (a percentage as large as 27.9% in Brazil in 2011 and 43.4% in Peru in 2018). Injuries and ill-defined diseases explain the remaining deaths.

Figure 2 (and Appendix Tables S9, S10, S11, and S12) report socio-economic inequality mortality ratios by sex and age group, according to whether the cause of death was a communicable, maternal or nutritional or non-communicable disease.

**Figure 2.**
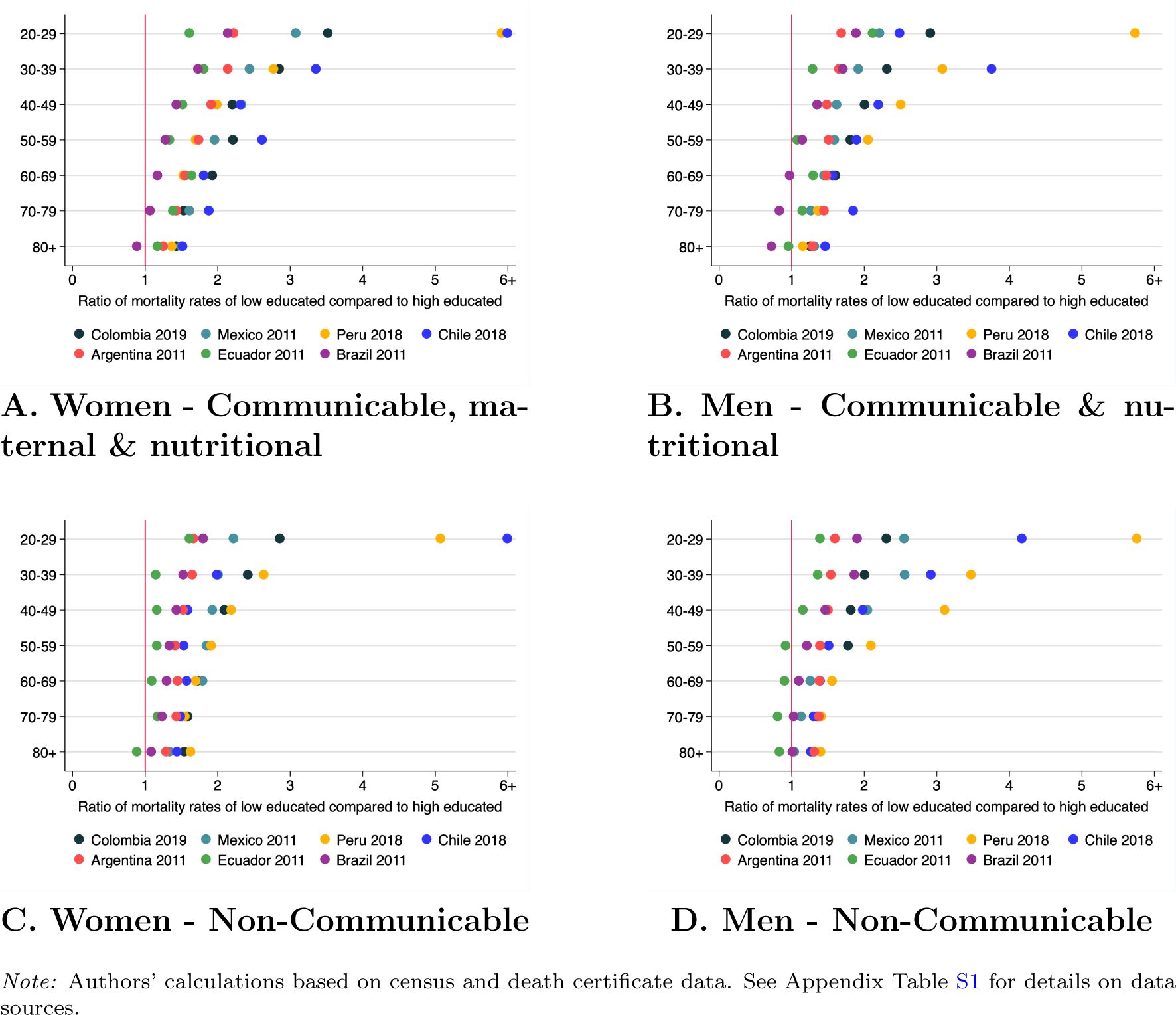
Socioeconomic inequality in mortality, by age, sex and cause (Communicable vs. Non-communicable)

Not surprisingly, the patterns of inequality by age and sex and across countries, particularly at younger ages, are still very significant, irrespective of the causes of death. For women, inequalities in mortality due to communicable, maternal and nutritional diseases are much larger than for non-communicable diseases: the unweighted median inequality ratio ranges from 3.08 to 1.37 for communicable, maternal and nutritional causes, while for non-communicable causes ranges from 2.22 to 1.34. The relation is more nuanced for men, with higher inequality for non-communicable diseases in early young groups (40-49 and younger) but, in general, higher mortality inequality due to communicable and nutritional diseases at older ages.

Appendix Tables S13, S14, S15, and S17 report socio-economic inequalities within communicable, maternal, and nutritional diseases. These socio-economic inequalities in mortality due to communicable diseases are larger than for maternal deaths. For example, in women aged between 20 and 29, the unweighted median inequality ratio for communicable diseases is 3.58, while for maternal deaths is 2.33. The mortality rates due to nutritional causes are near zero, except for older ages.

Appendix Tables S19, S21, and S23 report socio-economic inequalities within some main categories of non-communicable diseases (NCDs): cardiovascular, diabetes, and neoplasms. Socio-economic inequalities due to diabetes-related deaths are larger than for cardiovascular-related deaths for all age groups (both sexes) except for men in the age groups 20-29, 70-79, and 80+. Socio-economic inequalities on neoplasms-related deaths are the smallest of the three, with the unweighted median ratio across countries smaller than 2 for all age groups (for women ranging from 1.56 to 1.19 across age groups, and for men from 1.47 to 0.98).

#### 3.2.2 Socioeconomic Disparities in Mortality Rates by Injuries (Intentional and Unintentional)

Intentional and unintentional injuries represent 9.8 percent of all the deaths in our sample (see Appendix Table S6). Of these deaths, 45 percent are classified as intentional. Figure 3 and Tables S25, S26, S27, and S28 in the Appendix report (except for Chile that does not report intentional injuries as a primary cause of death) socio-economic inequality mortality ratios by sex and age group, according to whether the cause of death was intentional or unintentional injury.

**Figure 3.**
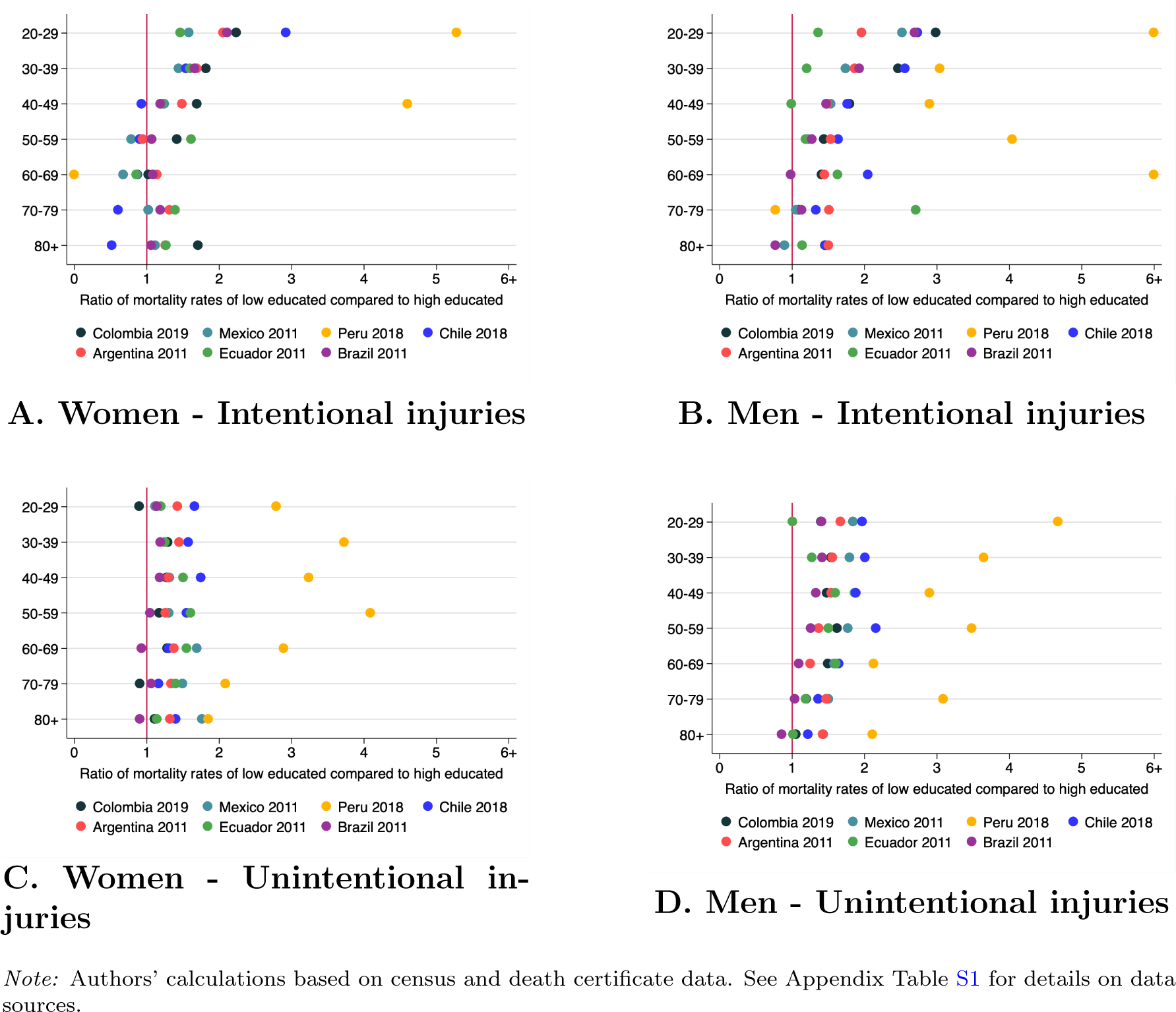
Socioeconomic inequality in mortality, by age, sex and cause (Intentional vs. Unintentional injuries)

The unweighted median mortality ratio from intentional injuries tends to decrease with age for both men and women. For instance, the unweighted mortality ratio for women is larger than 1.35 for the age group 40-49 and younger, while the ratio is smaller than 1.10 for the age groups 50-59 and 60-69. The relation between the unweighted mortality ratio and age is more nuanced for unintentional injuries.

Across both intentional and unintentional injuries, the socio-economic mortality inequality is higher for men than women: across all ages, the unweighted median mortality ratio is higher for men than women, except for intentional injuries in the age groups 70-79 and 80+

Within intentional injuries, Appendix Tables S29, S30 report suicide mortality rates and ratios while S31 and S32 report them for homicides. In each age group and sex, the homicide and suicide rates are higher amongst lower educated than higher educated, with a few exceptions, in homicides for women in the 60-69 age group and in suicides for women in the age groups 50-59 and 60-69. As in all-cause mortality, the unweighted mortality ratio for suicides and homicides also tends to decrease with age. For the age groups 20-29 and 30-39, the unweighted mortality ratio is higher for homicides than suicides, while the relation is nuanced for older age groups.

## 4 DISCUSSION

This study provides a comprehensive analysis of mortality inequalities across socioeconomic groups, age ranges, and sex in Latin America, offering crucial insights into the complex landscape of health disparities in the region that complements ongoing efforts to characterize life expectancy and mortality in Latin American cities.^11^ Our findings must be interpreted within the broader context of socio-economic, epidemiological, and health system changes that have occurred in Latin America over the past several decades, including an unprecedented surge in violence since the 2010s.^6,7,22,23,24^

Concurrent with a fall in income inequality,^24^ LAC has experienced a significant epidemiological transition, consisting of a broad shift from communicable, maternal, neonatal, and nutritional disorders towards non-communicable diseases and injuries between 1990 and 2010.^19^ The burden of disease has also shifted from children under 5 to reproductive age groups (15-49 years).

Our results show that socio-economic inequalities on mortality are more pronounced in younger age groups. This pattern persists across various causes of death, both for diseases and injuries. These findings suggest that despite overall improvements in income inequality and access to health services, significant challenges remain in addressing health disparities, particularly for younger populations.

The larger inequalities in younger age groups may reflect the persistent challenges in providing equitable access to preventive care and early intervention, especially for lower socioeconomic groups.

Whether inequalities are larger for men or women depends on the age group. For younger age groups (20-29, 30-39), inequalities tend to be larger amongst men than women. They are very similar for the 40-49 age group, while for those 50 or older, the inequalities tend to be larger amongst women than men.

The observed patterns in mortality from intentional injuries, including the higher rates among lower education groups, particularly before age 50, underscore the complex interplay between socioeconomic factors and violence that has also been shown in ecological studies.^25^ The varied sex patterns across countries (e.g., higher inequality for men in Argentina, Colombia, and Mexico at almost all age groups, but more nuanced patterns in Brazil and Ecuador) highlight the need for context-specific approaches to addressing violent mortality.

The hierarchy of socioeconomic inequalities across different non-violent causes of death (the largest for communicable diseases, followed by diabetes and cardiovascular diseases, and the smallest for neoplasms) provides valuable insights for health policy. The larger inequalities in communicable diseases and diabetes-related deaths suggest that these areas may benefit most from targeted interventions aimed at reducing health disparities.

Our findings reveal critical implications for regional policy, which needs to consider greater disparities among younger age groups. Additionally, a comprehensive approach that considers socioeconomic determinants of health and healthcare system factors is essential to address mortality inequalities effectively. Given the varied patterns of violent mortality across countries and socioeconomic groups, context-specific strategies for violence prevention are also necessary.

The growing burden of NCDs necessitates greater focus on equitable prevention and management strategies, particularly for diabetes and cardiovascular diseases. This shift is crucial as the region continues its epidemiological transition, with NCDs becoming increasingly prevalent across all socioeconomic groups. However, diverting resources needed to address NCDs away from efforts to treat and prevent communicable diseases could exacerbate existing inequalities in communicable diseases, which already show the greatest disparities. To reduce health inequalities during this transition, health systems will require more efficient ways to use existing resources, additional resources and increased policy attention.^26^

Continued health system reform remains a priority, with efforts to achieve universal health coverage needing further strengthening. Particular emphasis should be placed on reducing barriers to access for lower socioeconomic groups, ensuring that healthcare services are both accessible and affordable for all segments of the population.

Health production is influenced by a variety of interrelated individual and societal factors, including household resources, empowerment, information, and prevailing societal norms. Although expanding healthcare coverage and addressing barriers to accessing quality healthcare are essential steps in tackling health inequalities, these efforts may not be insufficient. Policies designed to reduce health disparities must go beyond the healthcare sector and address the broader social determinants.^27^

This study provides valuable insights but also has limitations. The cross-sectional nature of the data limits the study of how inequality of mortality rates changes over time. Future research should aim to establish how these inequalities evolved, particularly after the COVID-19 pandemic and recent Denge and Zika regional epidemics. Our analysis is also impaired by low-quality reporting in death certificate data, particularly missing information on education for some countries and under-reporting of deaths in others.

In conclusion, while Latin America has significantly reduced income inequality and improved overall health indicators, substantial mortality inequalities persist. Complex interactions between socioeconomic status, age, sex, and cause of death shape these inequalities. Addressing these disparities will require sustained, targeted efforts sensitive to the unique contexts of each country in the region.

## Data Availability

All data sources used in this study are publicly available and were obtained from official repositories. Census and death certificate data were accessed from the national statistical agencies of the participating countries. The specific sources and methods for data retrieval are detailed in the Methods section of the manuscript. Any additional information about the data can be provided upon reasonable request.

## Footnotes

1 The data quality criteria for death certificates include: completeness of mortality registration, ill-defined codes, and timeliness of reporting

