## Supplemental Appendix for "Socio-economic Disparities in Adult Mortality in Latin America*"

### 7 Appendix

#### 7.1 Appendix: sources of information and data harmonization

##### 7.1.1 Sources

We gather data from death certificates from every Latin American country that makes them publicly available, either as microdata or as tabulations required for our analysis.

We rely on population census data from each country to count the population by the required categories (age, sex, and educational attainment). We use the latest census publicly available for each country, either as microdata or as tabulations required for our analysis.

Table S1: Data Sources

| | Death certificates | | Population |
|---|---|---|---|
| | **Year** | **Education, by age, sex and cause of death** | **Education, by age and sex** |
| **Argentina** | 2011 | Tabulations provided by the Ministry of Health ( through a request for Public Information) | Online processing (08/09/2023) using REDATAM (www.redatam.indec.gob.ar). |
| **Brazil** | 2011 | Own tabulations from microdata from Instituto de Estudios para Políticas de Saúde. | Own estimations using IPUMS census 10% sample (www.ipums.org). |
| **Chile** | 2018 | Own calculation from death certificates microdata from Instituto Nacional de Estadísticas (INE) (through a request for public information) | Online processing (08/17/2023) using REDATAM (https://redatam-ine.ine.cl/redbin/RpWebEngine.exe/Portal?BASE=CENSO_2017&lang=esp) |
| **Colombia** | 2019 | Own calculation from death certificates microdata from Departamento Administrativo Nacional de Estadística (DANE) (https://microdatos.dane.gov.co/index.php/catalog/DEM-Microdatos) | Online processing (07/17/2023) using REDATAM (https://redatam-ine.ine.cl/redbin/RpWebEngine.exe/Portal?BASE=CENSO_2017&lang=esp) |
| **Ecuador** | 2011 | Own calculation from death certificates microdata from INEC (http://redatam.inec.gob.ec/cgibin/RpWebEngine.exe/PortalAction?BASE=CPV2010) | Online processing (12/26/2023) using REDATAM (http://redatam.inec.gob.ec/cgibin/RpWebEngine.exe/PortalAction?BASE=CPV2010) |
| **Mexico** | 2011 | Own calculation from death certificates microdata from INEGI (https://www.inegi.org.mx/programas/mortalidad/?ps=Microdatos) | Online processing (07/20/2023) using Subsistema de Información Demográfica y Social (https://www.inegi.org.mx/programas/ccpv/2010/default.html#Tabulados) |
| **Peru** | 2018 | Own calculation from death certificates microdata from Sistema Informático Nacional de Defunciones (SINADEF) (https://www.datosabiertos.gob.pe/dataset/informacin-de-fallecidos-del-sistema-informtico-nacional-de-defunciones-sinadef-ministerio) | Online processing (07/17/2023) using REDATAM (https://censo2017.inei.gob.pe) |

##### 7.1.2 Data harmonization

Data are harmonized to ensure comparability across countries and between death certificates and census data within countries.

##### 7.1.3 Harmonization of education categories

Table S2 presents a full description of the harmonization of education variables.

#### Table S2: Harmonization of education categories

| | Death Certificates | | | Census | | |
|---|---|---|---|---|---|---|
| | **Q** | **Low** | **High** | **Q** | **Low** | **High** |
| Argentina 2011 | Highest level of education attained by the deceased | Nunca asistió; Primario incompleto; Primario Completo; Secundario incompleto; Ciclos EGB (1 y 2) incompleto; Ciclos EGB (1 y 2) completo; Ciclo EGB 3 incompleto; Ciclo EGB 3 completo; Polimodal incompleto | Secundario completo; Polimodal completo; Superior o Universitario incompleto; Superior o universitario completo | Highest level of education attained | Primario incompleto; Primario completo; Secundario incompleto; EGB incompleto; EGB completo; Polimodal incompleto | Secundario completo; Polimodal completo; Superior no universitario incompleto; Universitario incompleto; Superior no universitario completo; Universitario completo; Post universitario incompleto; Post universitario completo |
| Brazil 2011 | Schooling years of the deceased | No schooling; From 1 to 3 years; From 4 to 7 years | From 8 to 11 years; 12 years or more | Highest level of education attained | None or never attended school; Attending day-care center; Attending preschool; Attending child literacy course; Attending adult literacy course; Attended adult literacy course; Attended day-care center, pre-school, or child literacy course; Attending first grade of primary school; Primary - Grade 1; Primary - Grade 2; Primary - Grade 3; Primary - Grade 4; Primary - Grade 5; Primary - Grade 6; Primary - Grade 7; Primary - Grade unspecified | Primary - Grade 8; Secondary - Grade 1; Secondary - Grade 2; Secondary - Grade 3; Secondary - Grade unspecified; Undergraduate program - Attending; Undergraduate program - Attended; Undergraduate program - Completed; Masters, attending; Masters, attended; Doctorate, attending; Doctorate, attended; Postgraduate specialization, attending; Postgraduate specialization, attended |
| Chile 2018 | Highest level of education attained | Sin instrucción, Básico o primaria; Secundaria 1; Secundaria 2 | Secundaria 3; Secundaria 4; Secundaria 5; Secundaria 6; Secundaria 7; Medio; Superior | Schooling years | Hasta 9 años de estudio | De 10 a 12 años de estudio; 13 años y más de estudio |
| Colombia 2019 | Highest level of education attained by the deceased | Ninguno; Preescolar; Básica primaria; Básica secundaria | Media académica o clásica; Media técnica; Normalista; Técnica profesional; Tecnológica; Profesional; Especialización; Maestría; Doctorado | Highest level of education attained | Ninguno; Preescolar (Prejardin); Preescolar (Jardin); Preescolar (Transición); Básica primaria 1; Básica primaria 2; Básica primaria 3; Básica primaria 4; Básica primaria 5; Básica secundaria 6; Básica secundaria 7; Básica secundaria 8; Básica secundaria 9; Media academica o clasica 10; Media tecnica 10; Normalista 10; Normalista 11; Normalista 12 | Media academica o clasica 11; Media tecnica 11; Normalista 13; Técnica profesional 1 año; Técnica profesional 2 años; Técnica profesional 3 años; Tecnológica 1 año; Tecnológica 2 años; Tecnológica 3 años; Universitario 1 año; Universitario 2 años; Universitario 3 años; Universitario 4 años; Universitario 5 años; Universitario 6 años; Especialización 1 año; Especialización 2 años; Maestria 1 año; Maestria 2 años; Maestria 3 años; Doctorado 1 año; Doctorado 2 años; Doctorado 3 años; Doctorado 4 años; Doctorado 5 años; Doctorado 6 años |
| Ecuador 2011 | Highest level of education attained by the deceased | Ninguno; Centro de alfabetización; Primario; Educación básica | Secundaria; Educación media; Ciclo post bachillerato; Superior; Postgrado | Highest level of education attained | Preescolar; Primario; Educación Básica; Centro de Alfabetización/(EBA); Educación Media 1 año; Educación Media 2 años; Educación Media año no especificado; Secundario 1 año; Secundario 2 años; Secundario 3 años; Secundario 4 años; Secundario 5 años; Secundario año no especificado | Secundario 6 años; Educación Media 3 años; Ciclo Postbachillerato; Superior 1 año; Superior 2 años; Superior 3 años; Superior 4 años; Superior 5 años; Superior 6 años; Superior 7 años; Superior 8 años; Superior año no especificado; Postgrado |
| Mexico 2011 | Highest level of education attained by the deceased | No aplica a menores de 6 años; Sin escolaridad; Primaria incompleta; Primaria completa; Secundaria incompleta; Secundaria completa | Bachillerato o preparatoria; Profesional | Highest level of education attained | Sin escolaridad; Educacion basica - Preescolar; Educacion basica - Primaria; Educacion basica - Secundaria Incompleta; Educacion basica - Secundaria Completa; Educacion basica - Secundaria No especificado | Estudios técnicos o comerciales con primaria terminada; Educación media superior; Educación superior |
| Peru 2018 | Highest level of education attained by the deceased | Ningún nivel / Iletrado; Inicial / Preescolar; Primaria incompleta; Primaria completa; Secundaria incompleta | Secundaria completa; Superior no universitaria incompleta; Superior no universitaria completo; Superior universitaria incompleta; Superior universitaria completa | Highest level of education attained | Inicial; Basica especial; Primaria | Secundaria; Superior no universitaria incompleta; Superior no universitaria completa; Superior universitaria incompleta; Superior universitaria completa; Maestria/doctorado |

Table S3: Share of population with lower level of education by age, gender and country

| | | Argentina 2011 | Brazil 2011 | Chile 2018 | Colombia 2019 | Ecuador 2011 | Mexico 2011 | Peru 2018 |
|---|---|---|---|---|---|---|---|---|
| 20-29 | Women | 39% | 32% | 8% | 21% | 43% | 50% | 12% |
| | Men | 49% | 40% | 11% | 29% | 45% | 52% | 9% |
| 30-39 | Women | 43% | 46% | 13% | 30% | 52% | 62% | 22% |
| | Men | 51% | 53% | 15% | 36% | 54% | 61% | 15% |
| 40-49 | Women | 50% | 57% | 24% | 45% | 56% | 65% | 30% |
| | Men | 57% | 62% | 26% | 50% | 57% | 62% | 21% |
| 50-59 | Women | 57% | 66% | 38% | 58% | 66% | 74% | 40% |
| | Men | 63% | 68% | 38% | 59% | 65% | 69% | 28% |
| 60-69 | Women | 66% | 78% | 51% | 70% | 78% | 83% | 54% |
| | Men | 69% | 77% | 49% | 69% | 76% | 80% | 38% |
| 70-79 | Women | 76% | 86% | 66% | 82% | 85% | 90% | 71% |
| | Men | 74% | 84% | 62% | 79% | 84% | 89% | 56% |
| 80+ | Women | 82% | 89% | 72% | 89% | 89% | 93% | 77% |
| | Men | 77% | 87% | 68% | 86% | 88% | 92% | 68% |

*Note:* Authors' calculations based on census and death certificate data. See Appendix Table S1 for details on data sources.

###### 7.1.4 Harmonization of cause of death

All death certificates used the International Classification of Diseases (ICD-10) codes to classify causes of death. Table (S4) shows the classification of the ICD-10 codes used to divide the causes of death into major groups. We used the groups of the *WHO mortality database.*

Table S4: ICD-10 codes used to identify groups of diseases

| Disease | ICD-10 Codes |
|---|---|
| Communicable-Maternal-Nutritional | A00-A99, B00-B99, D50-D53, E00-E02, E40-E46, E50, E51-E64, G00-G05, G14, H65-H66, J00-J22, L00-L08, N70-N73, O00-O99, P23, P37, U04, U07, U09, U10, Y95 |
| Communicable | A00-A99, B00-B99, G00-G05, G14, H65-H66, J00-J22, L00-L08, N70-N73, P23, P37, U04, U07, U09, U10, Y95 |
| Maternal | O00-O99 |
| Nutritional | D50-D53, E00-E02, E40-E46, E50, E51-E64 |
| Non-Communicable | C00-C97, D00-D48, D55-D64, D65-D89, E03-E07, E10-E34, E65-E88, F00-F99, G06-G13, G15-G99, H00-H61, H68-H93, I00-I99, J30-J99, K00-K93, L09-L99, M00-M99, N00-N64, N75-N98, Q00-Q99, R95, X41, X42, X44, X45 |
| Cardiovascular diseases | I00-I99 |
| Neoplasms | C00-C97, D00-D48 |
| Diabetes | N18, N00, N03, N05, N07, N08, N29, N17, N19, E10, E11, E13, E14 |
| Intentional injuries | X60-X99, Y00-Y09, Y35-Y36, Y87 |
| Homicide | X85-X99, Y00-Y09 |
| Suicide | X60-X84 |
| Unintentional Injuries | S00-S99, T00-T99, U12, V01-V99, W00-W99, X00-X40, X43, X46-X59, Y10-Y34, Y40-Y86, Y85, Y86, Y88, Y89, Y90, Y91 |

###### 7.1.5 Missing information

Table S5 presents the percentage of valid (non-missing) data for each variable —sex, age, educational attainment, and cause of death—across the census and death certificate datasets by country. Reporting for sex and age showed no significant issues, with at least 99% coverage across all datasets. However, Peru is the only country where missing values for the cause of death are substantial (28%). Since this issue cannot be resolved, these deaths were excluded from the cause-of-death analysis. Additionally, the Chilean death certificate dataset lacked information on intentional causes of death, such as suicide and homicide.

To deal with missing information for educational attainment, we assume that the distribution of educational attainment among individuals with missing educational attainment is the same as the distribution observed in the census for individuals of the same age and sex.

Guatemala also includes educational attainment in the death certificates, but it was omitted from the analysis because the education attainment is missing for 23% of the population listed in the census.

Table S5: Percentage of valid data for each variable by source (census and death certificates) and by country

| Country | Census data | | | Deaths certificates data | | | |
|---|---|---|---|---|---|---|---|
| | Age | Sex | Education | Age | Sex | Education | Cause of death |
| Argentina | 98.9% | 100.0% | 96.6% | 99.7% | 99.9% | 51.8% | 100.0% |
| Brazil | 100.0% | 100.0% | 100.0% | 99.6% | 100.0% | 75.1% | 99.3% |
| Chile | 100.0% | 100.0% | 97.3% | 100.0% | 100.0% | 99.9% | 100.0% |
| Colombia | 100.0% | 100.0% | 98.4% | 100.0% | 100.0% | 81.0% | 100.0% |
| Ecuador | 100.0% | 100.0% | 96.4% | 99.8% | 100.0% | 94.0% | 100.0% |
| Mexico | 100.0% | 100.0% | 99.4% | 99.2% | 100.0% | 95.0% | 100.0% |
| Peru | 100.0% | 100.0% | 100.0% | 99.8% | 100.0% | 79.8% | 72.8% |

*Note:* Authors' calculations based on census and death certificate data. See Table S1 for details on data sources.

#### 7.2 Appendix: Additional Results

Prevalence of diseases

Table S6: Percentage of each disease group per country

| | Argentina 2011 | Brasil 2011 | Chile 2018 | Colombia 2019 | Ecuador 2011 | Mexico 2011 | Peru 2018 | All |
|---|---|---|---|---|---|---|---|---|
| Communicable, maternal & nutritional | 11.68 | 27.87 | 7.37 | 20.2 | 9.91 | 7.33 | 43.38 | 19.67 |
| Non Communicable | 74.41 | 55.59 | 83.4 | 69.87 | 66.56 | 79.44 | 52.38 | 66.04 |
| Intentional injuries | 1.3 | 4.77 | 2.06 | 6.09 | 4.62 | 5.07 | 0.18 | 4.26 |
| Unintentional injuries | 4.12 | 5.21 | 4.78 | 3.16 | 9.36 | 6.24 | 2.64 | 5.1 |
| Ill-defined diseases | 8.5 | 6.56 | 2.39 | 0.67 | 9.56 | 1.92 | 1.43 | 4.93 |

*Note:* Authors' calculations based on census and death certificate data. See Appendix Table S1 for details on data sources.

All causes

Table S7: Mortality rates by sex, age and education - All causes

| Country | Education | 20-29 | 30-39 | 40-49 | 50-59 | 60-69 | 70-79 | 80+ |
|---|---|---|---|---|---|---|---|---|
| | | | | Women | | | | |
| Argentina 2011 | Low | 0.74 | 1.24 | 2.63 | 5.82 | 13.31 | 31.87 | 121.60 |
| | High | 0.42 | 0.72 | 1.69 | 4.06 | 9.10 | 22.35 | 94.85 |
| Brazil 2011 | Low | 0.94 | 1.43 | 2.89 | 6.07 | 13.08 | 31.25 | 98.97 |
| | High | 0.51 | 0.90 | 2.01 | 4.55 | 10.26 | 25.99 | 95.31 |
| Chile 2018 | Low | 1.27 | 1.03 | 1.89 | 4.08 | 10.04 | 24.23 | 95.17 |
| | High | 0.28 | 0.53 | 1.16 | 2.62 | 6.39 | 16.07 | 65.33 |
| Colombia 2019 | Low | 1.24 | 1.61 | 2.48 | 4.44 | 10.07 | 24.44 | 91.06 |
| | High | 0.48 | 0.69 | 1.21 | 2.30 | 5.76 | 15.60 | 60.22 |
| Ecuador 2011 | Low | 0.88 | 1.23 | 2.35 | 5.04 | 11.43 | 32.69 | 136.04 |
| | High | 0.57 | 0.91 | 1.77 | 3.92 | 9.40 | 25.12 | 129.01 |
| Mexico 2011 | Low | 0.42 | 0.59 | 1.31 | 3.24 | 7.43 | 16.68 | 52.27 |
| | High | 0.22 | 0.31 | 0.69 | 1.78 | 4.19 | 10.87 | 37.31 |
| Peru 2018 | Low | 1.56 | 1.43 | 2.37 | 4.36 | 8.63 | 20.01 | 69.04 |
| | High | 0.37 | 0.56 | 1.15 | 2.54 | 5.49 | 13.75 | 49.71 |
| | | | | Men | | | | |
| Argentina 2011 | Low | 1.98 | 2.19 | 4.33 | 10.58 | 25.29 | 57.75 | 155.37 |
| | High | 1.16 | 1.37 | 2.83 | 7.48 | 18.09 | 41.33 | 117.92 |
| Brazil 2011 | Low | 3.75 | 3.92 | 5.87 | 11.12 | 21.52 | 45.98 | 116.73 |
| | High | 1.83 | 2.24 | 4.07 | 9.09 | 19.83 | 46.42 | 126.35 |
| Chile 2018 | Low | 2.34 | 3.16 | 4.03 | 7.59 | 16.56 | 38.90 | 116.03 |
| | High | 0.87 | 1.23 | 2.07 | 4.76 | 11.59 | 29.15 | 88.96 |
| Colombia 2019 | Low | 4.27 | 3.81 | 3.95 | 6.88 | 16.06 | 36.78 | 107.91 |
| | High | 1.72 | 1.78 | 2.20 | 3.97 | 10.30 | 27.15 | 83.26 |
| Ecuador 2011 | Low | 2.80 | 3.01 | 3.98 | 6.93 | 14.77 | 39.33 | 135.82 |
| | High | 2.18 | 2.29 | 3.02 | 6.38 | 13.73 | 41.46 | 143.01 |
| Mexico 2011 | Low | 1.64 | 1.96 | 2.94 | 5.21 | 10.09 | 21.03 | 56.43 |
| | High | 0.72 | 0.97 | 1.55 | 3.43 | 7.90 | 18.16 | 51.23 |
| Peru 2018 | Low | 4.41 | 3.52 | 4.51 | 7.01 | 11.75 | 26.10 | 78.08 |
| | High | 0.87 | 1.15 | 1.79 | 3.63 | 7.93 | 19.43 | 64.00 |

*Note:* Authors' calculations based on census and death certificate data. See Appendix Table S1 for details on data sources.

Table S8: Mortality rates ratios by sex, age and education - All causes

| Country | 20-29 | 30-39 | 40-49 | 50-59 | 60-69 | 70-79 | 80+ |
|---|---|---|---|---|---|---|---|
| Women | | | | | | | |
| Argentina 2011 | 1.77 | 1.72 | 1.56 | 1.43 | 1.46 | 1.43 | 1.28 |
| Brazil 2011 | 1.83 | 1.59 | 1.44 | 1.33 | 1.27 | 1.20 | 1.04 |
| Chile 2018 | 4.54 | 1.96 | 1.63 | 1.56 | 1.57 | 1.51 | 1.46 |
| Colombia 2019 | 2.60 | 2.35 | 2.05 | 1.93 | 1.75 | 1.57 | 1.51 |
| Ecuador 2011 | 1.55 | 1.36 | 1.33 | 1.28 | 1.22 | 1.30 | 1.05 |
| Mexico 2011 | 1.93 | 1.89 | 1.89 | 1.83 | 1.78 | 1.53 | 1.40 |
| Peru 2018 | 4.29 | 2.55 | 2.05 | 1.72 | 1.57 | 1.45 | 1.39 |
| Median | 1.93 | 1.89 | 1.63 | 1.56 | 1.57 | 1.45 | 1.39 |
| Max/Min | 2.93 | 1.88 | 1.55 | 1.50 | 1.46 | 1.30 | 1.46 |
| Men | | | | | | | |
| Argentina 2011 | 1.72 | 1.60 | 1.53 | 1.41 | 1.40 | 1.40 | 1.32 |
| Brazil 2011 | 2.05 | 1.75 | 1.44 | 1.22 | 1.09 | 0.99 | 0.92 |
| Chile 2018 | 2.69 | 2.57 | 1.95 | 1.60 | 1.43 | 1.33 | 1.30 |
| Colombia 2019 | 2.48 | 2.15 | 1.80 | 1.73 | 1.56 | 1.36 | 1.30 |
| Ecuador 2011 | 1.29 | 1.31 | 1.32 | 1.09 | 1.08 | 0.95 | 0.95 |
| Mexico 2011 | 2.27 | 2.03 | 1.90 | 1.52 | 1.28 | 1.16 | 1.10 |
| Peru 2018 | 5.06 | 3.05 | 2.52 | 1.93 | 1.48 | 1.34 | 1.22 |
| Median | 2.27 | 2.03 | 1.80 | 1.52 | 1.40 | 1.33 | 1.22 |
| Max/Min | 3.93 | 2.32 | 1.92 | 1.78 | 1.45 | 1.47 | 1.43 |

*Note:* Authors' calculations based on census and death certificate data. See Appendix Table S1 for details on data sources.

Communicable, maternal & nutritional causes

Table S9: Mortality rates by sex, age and education - Communicable, maternal & nutritional causes

| Country | Education | 20-29 | 30-39 | 40-49 | 50-59 | 60-69 | 70-79 | 80+ |
|---|---|---|---|---|---|---|---|---|
| | | | | Women | | | | |
| Argentina 2011 | Low | 0.15 | 0.26 | 0.33 | 0.50 | 1.23 | 3.56 | 17.00 |
| | High | 0.07 | 0.12 | 0.17 | 0.29 | 0.79 | 2.49 | 13.61 |
| Brazil 2011 | Low | 0.31 | 0.47 | 0.73 | 1.50 | 3.51 | 9.47 | 34.18 |
| | High | 0.14 | 0.27 | 0.51 | 1.17 | 2.98 | 8.82 | 38.56 |
| Chile 2018 | Low | 0.18 | 0.15 | 0.15 | 0.24 | 0.52 | 1.53 | 9.25 |
| | High | 0.02 | 0.04 | 0.06 | 0.09 | 0.29 | 0.81 | 6.10 |
| Colombia 2019 | Low | 0.41 | 0.46 | 0.53 | 0.88 | 1.94 | 4.96 | 20.47 |
| | High | 0.12 | 0.16 | 0.24 | 0.40 | 1.01 | 3.23 | 14.28 |
| Ecuador 2011 | Low | 0.20 | 0.30 | 0.25 | 0.31 | 0.69 | 2.42 | 17.76 |
| | High | 0.13 | 0.16 | 0.17 | 0.23 | 0.42 | 1.74 | 15.16 |
| Mexico 2011 | Low | 0.08 | 0.09 | 0.10 | 0.17 | 0.37 | 0.93 | 4.77 |
| | High | 0.03 | 0.04 | 0.04 | 0.09 | 0.24 | 0.58 | 3.17 |
| Peru 2018 | Low | 0.45 | 0.36 | 0.53 | 1.09 | 2.39 | 6.61 | 26.86 |
| | High | 0.08 | 0.13 | 0.27 | 0.64 | 1.56 | 4.61 | 19.58 |
| | | | | Men | | | | |
| Argentina 2011 | Low | 0.12 | 0.26 | 0.52 | 0.89 | 2.12 | 6.12 | 22.06 |
| | High | 0.07 | 0.16 | 0.35 | 0.59 | 1.43 | 4.23 | 17.11 |
| Brazil 2011 | Low | 0.35 | 0.69 | 1.24 | 2.55 | 5.31 | 13.20 | 37.83 |
| | High | 0.18 | 0.40 | 0.91 | 2.21 | 5.43 | 15.77 | 52.21 |
| Chile 2018 | Low | 0.12 | 0.31 | 0.35 | 0.50 | 0.79 | 2.37 | 11.56 |
| | High | 0.05 | 0.08 | 0.16 | 0.26 | 0.50 | 1.28 | 7.89 |
| Colombia 2019 | Low | 0.47 | 0.59 | 0.74 | 1.25 | 3.11 | 7.41 | 23.93 |
| | High | 0.16 | 0.26 | 0.37 | 0.69 | 1.93 | 5.39 | 18.94 |
| Ecuador 2011 | Low | 0.31 | 0.39 | 0.34 | 0.53 | 0.96 | 3.25 | 15.58 |
| | High | 0.15 | 0.30 | 0.25 | 0.49 | 0.74 | 2.83 | 16.28 |
| Mexico 2011 | Low | 0.11 | 0.19 | 0.24 | 0.30 | 0.51 | 1.21 | 5.50 |
| | High | 0.05 | 0.10 | 0.15 | 0.19 | 0.35 | 0.95 | 4.18 |
| Peru 2018 | Low | 0.81 | 0.74 | 0.98 | 1.73 | 3.28 | 8.18 | 29.91 |
| | High | 0.14 | 0.24 | 0.39 | 0.84 | 2.18 | 5.99 | 25.91 |

*Note:* Authors' calculations based on census and death certificate data. See Appendix Table S1 for details on data sources.

Table S10: Mortality rates ratios by sex, age and education - Communicable, maternal & nutritional causes

| Country | 20-29 | 30-39 | 40-49 | 50-59 | 60-69 | 70-79 | 80+ |
|---|---|---|---|---|---|---|---|
| Women | | | | | | | |
| Argentina 2011 | 2.22 | 2.14 | 1.91 | 1.74 | 1.55 | 1.43 | 1.25 |
| Brazil 2011 | 2.14 | 1.74 | 1.44 | 1.29 | 1.18 | 1.07 | 0.89 |
| Chile 2018 | 9.40 | 3.36 | 2.32 | 2.61 | 1.81 | 1.89 | 1.52 |
| Colombia 2019 | 3.52 | 2.86 | 2.20 | 2.21 | 1.93 | 1.54 | 1.43 |
| Ecuador 2011 | 1.62 | 1.81 | 1.52 | 1.34 | 1.65 | 1.39 | 1.17 |
| Mexico 2011 | 3.08 | 2.45 | 2.33 | 1.96 | 1.56 | 1.62 | 1.51 |
| Peru 2018 | 5.92 | 2.77 | 1.99 | 1.70 | 1.53 | 1.43 | 1.37 |
| Median | 3.08 | 2.45 | 1.99 | 1.74 | 1.56 | 1.43 | 1.37 |
| Max/Min | 5.81 | 1.93 | 1.63 | 2.03 | 1.64 | 1.75 | 1.71 |
| Men | | | | | | | |
| Argentina 2011 | 1.69 | 1.65 | 1.49 | 1.51 | 1.48 | 1.45 | 1.29 |
| Brazil 2011 | 1.89 | 1.71 | 1.35 | 1.15 | 0.98 | 0.84 | 0.72 |
| Chile 2018 | 2.49 | 3.76 | 2.20 | 1.90 | 1.57 | 1.85 | 1.46 |
| Colombia 2019 | 2.91 | 2.31 | 2.01 | 1.81 | 1.61 | 1.37 | 1.26 |
| Ecuador 2011 | 2.12 | 1.30 | 1.35 | 1.08 | 1.30 | 1.15 | 0.96 |
| Mexico 2011 | 2.22 | 1.92 | 1.63 | 1.59 | 1.45 | 1.27 | 1.31 |
| Peru 2018 | 5.74 | 3.08 | 2.51 | 2.06 | 1.50 | 1.37 | 1.15 |
| Median | 2.22 | 1.92 | 1.63 | 1.59 | 1.48 | 1.37 | 1.26 |
| Max/Min | 3.40 | 2.90 | 1.85 | 1.90 | 1.64 | 2.21 | 2.02 |

*Note:* Authors' calculations based on census and death certificate data. See Appendix Table S1 for details on data sources.

Non-Communicable causes

Table S11: Mortality rates by sex, age and education - Non Communicable causes

| Country | Education | 20-29 | 30-39 | 40-49 | 50-59 | 60-69 | 70-79 | 80+ |
|---|---|---|---|---|---|---|---|---|
| | | | | Women | | | | |
| Argentina 2011 | Low | 0.31 | 0.72 | 1.93 | 4.71 | 10.84 | 25.09 | 90.17 |
| | High | 0.19 | 0.44 | 1.26 | 3.32 | 7.46 | 17.47 | 69.98 |
| Brazil 2011 | Low | 0.29 | 0.64 | 1.77 | 4.02 | 8.63 | 19.52 | 55.37 |
| | High | 0.16 | 0.42 | 1.23 | 3.01 | 6.62 | 15.74 | 50.80 |
| Chile 2018 | Low | 0.77 | 0.66 | 1.48 | 3.60 | 9.19 | 21.88 | 80.16 |
| | High | 0.11 | 0.33 | 0.93 | 2.35 | 5.82 | 14.69 | 55.64 |
| Colombia 2019 | Low | 0.53 | 0.92 | 1.76 | 3.41 | 7.96 | 19.18 | 69.58 |
| | High | 0.19 | 0.38 | 0.84 | 1.79 | 4.61 | 12.06 | 45.14 |
| Ecuador 2011 | Low | 0.33 | 0.61 | 1.59 | 3.96 | 9.26 | 25.78 | 95.24 |
| | High | 0.21 | 0.53 | 1.37 | 3.41 | 8.48 | 21.95 | 106.80 |
| Mexico 2011 | Low | 0.21 | 0.39 | 1.09 | 2.93 | 6.86 | 15.22 | 44.01 |
| | High | 0.09 | 0.20 | 0.57 | 1.58 | 3.82 | 10.02 | 32.92 |
| Peru 2018 | Low | 0.57 | 0.54 | 1.05 | 2.03 | 3.96 | 8.37 | 26.00 |
| | High | 0.11 | 0.20 | 0.48 | 1.06 | 2.33 | 5.39 | 15.93 |
| | | | | Men | | | | |
| Argentina 2011 | Low | 0.38 | 0.78 | 2.61 | 7.96 | 20.15 | 45.25 | 116.01 |
| | High | 0.24 | 0.50 | 1.73 | 5.70 | 14.54 | 32.75 | 88.24 |
| Brazil 2011 | Low | 0.46 | 1.05 | 2.72 | 6.50 | 13.65 | 28.52 | 67.03 |
| | High | 0.24 | 0.56 | 1.85 | 5.37 | 12.36 | 27.61 | 66.23 |
| Chile 2018 | Low | 0.74 | 1.27 | 2.46 | 5.73 | 14.29 | 34.48 | 97.99 |
| | High | 0.18 | 0.43 | 1.24 | 3.79 | 10.24 | 26.33 | 77.10 |
| Colombia 2019 | Low | 0.71 | 0.97 | 1.77 | 4.52 | 11.89 | 28.27 | 82.15 |
| | High | 0.31 | 0.49 | 0.98 | 2.54 | 7.62 | 20.82 | 62.89 |
| Ecuador 2011 | Low | 0.46 | 0.76 | 1.76 | 4.24 | 10.62 | 28.69 | 96.76 |
| | High | 0.33 | 0.56 | 1.52 | 4.61 | 11.71 | 35.48 | 116.36 |
| Mexico 2011 | Low | 0.37 | 0.76 | 1.84 | 4.14 | 8.76 | 18.67 | 46.92 |
| | High | 0.14 | 0.29 | 0.90 | 2.74 | 6.96 | 16.51 | 45.10 |
| Peru 2018 | Low | 1.02 | 0.99 | 1.62 | 2.85 | 4.84 | 11.05 | 29.63 |
| | High | 0.18 | 0.29 | 0.52 | 1.36 | 3.10 | 7.83 | 21.17 |

*Note:* Authors' calculations based on census and death certificate data. See Appendix Table S1 for details on data sources.

Table S12: Mortality rates ratios by sex, age and education - Non Communicable causes

| Country | 20-29 | 30-39 | 40-49 | 50-59 | 60-69 | 70-79 | 80+ |
|---|---|---|---|---|---|---|---|
| Women | | | | | | | |
| Argentina 2011 | 1.67 | 1.65 | 1.53 | 1.42 | 1.45 | 1.44 | 1.29 |
| Brazil 2011 | 1.81 | 1.53 | 1.44 | 1.34 | 1.30 | 1.24 | 1.09 |
| Chile 2018 | 7.15 | 1.99 | 1.59 | 1.53 | 1.58 | 1.49 | 1.44 |
| Colombia 2019 | 2.86 | 2.42 | 2.09 | 1.90 | 1.73 | 1.59 | 1.54 |
| Ecuador 2011 | 1.62 | 1.15 | 1.16 | 1.16 | 1.09 | 1.17 | 0.89 |
| Mexico 2011 | 2.22 | 2.01 | 1.93 | 1.85 | 1.80 | 1.52 | 1.34 |
| Peru 2018 | 5.08 | 2.64 | 2.19 | 1.91 | 1.70 | 1.55 | 1.63 |
| Median | 2.22 | 1.99 | 1.59 | 1.53 | 1.58 | 1.49 | 1.34 |
| Max/Min | 4.42 | 2.30 | 1.88 | 1.64 | 1.64 | 1.35 | 1.83 |
| Men | | | | | | | |
| Argentina 2011 | 1.60 | 1.54 | 1.51 | 1.40 | 1.39 | 1.38 | 1.31 |
| Brazil 2011 | 1.91 | 1.87 | 1.47 | 1.21 | 1.10 | 1.03 | 1.01 |
| Chile 2018 | 4.17 | 2.93 | 1.99 | 1.51 | 1.40 | 1.31 | 1.27 |
| Colombia 2019 | 2.30 | 2.01 | 1.82 | 1.78 | 1.56 | 1.36 | 1.31 |
| Ecuador 2011 | 1.39 | 1.36 | 1.16 | 0.92 | 0.91 | 0.81 | 0.83 |
| Mexico 2011 | 2.55 | 2.56 | 2.05 | 1.51 | 1.26 | 1.13 | 1.04 |
| Peru 2018 | 5.76 | 3.47 | 3.11 | 2.10 | 1.56 | 1.41 | 1.40 |
| Median | 2.30 | 2.01 | 1.82 | 1.51 | 1.39 | 1.31 | 1.27 |
| Max/Min | 4.13 | 2.55 | 2.69 | 2.28 | 1.72 | 1.74 | 1.68 |

*Note:* Authors' calculations based on census and death certificate data. See Appendix Table S1 for details on data sources.

#### Communicable

Figure S1: Communicable

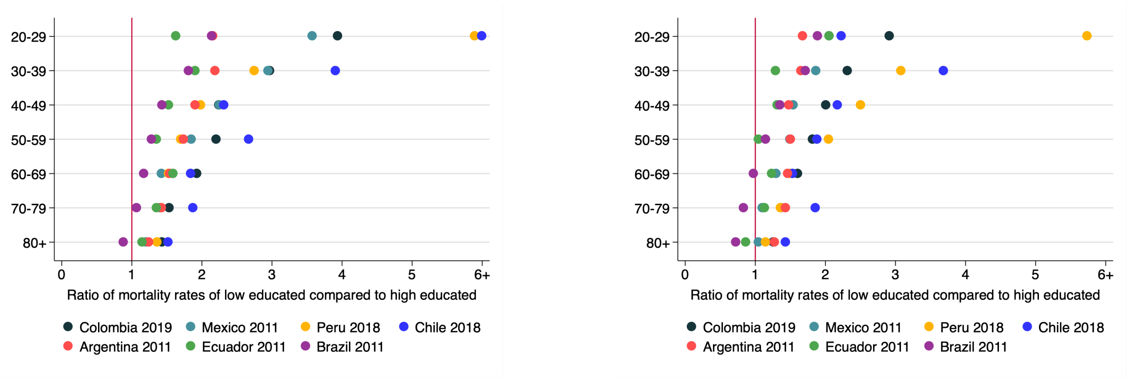

Women Men

*Note:* Authors' calculations based on census and death certificate data. See Appendix Table S1 for details on data sources.

Table S13: Mortality rates by sex, age and education - Communicable

| Country | Education | 20-29 | 30-39 | 40-49 | 50-59 | 60-69 | 70-79 | 80+ |
|---|---|---|---|---|---|---|---|---|
| | | | | Women | | | | |
| Argentina 2011 | Low | 0.09 | 0.19 | 0.30 | 0.49 | 1.21 | 3.47 | 16.46 |
| | High | 0.04 | 0.09 | 0.16 | 0.28 | 0.78 | 2.43 | 13.21 |
| Brazil 2011 | Low | 0.24 | 0.42 | 0.72 | 1.50 | 3.51 | 9.46 | 34.11 |
| | High | 0.11 | 0.23 | 0.50 | 1.17 | 2.98 | 8.82 | 38.53 |
| Chile 2018 | Low | 0.15 | 0.11 | 0.14 | 0.23 | 0.50 | 1.44 | 8.59 |
| | High | 0.01 | 0.03 | 0.06 | 0.09 | 0.27 | 0.77 | 5.64 |
| Colombia 2019 | Low | 0.29 | 0.36 | 0.51 | 0.88 | 1.94 | 4.96 | 20.46 |
| | High | 0.07 | 0.12 | 0.23 | 0.40 | 1.01 | 3.23 | 14.28 |
| Ecuador 2011 | Low | 0.11 | 0.17 | 0.21 | 0.29 | 0.64 | 2.18 | 14.91 |
| | High | 0.07 | 0.09 | 0.14 | 0.21 | 0.40 | 1.61 | 12.92 |
| Mexico 2011 | Low | 0.04 | 0.06 | 0.09 | 0.16 | 0.32 | 0.73 | 3.05 |
| | High | 0.01 | 0.02 | 0.04 | 0.08 | 0.22 | 0.53 | 2.52 |
| Peru 2018 | Low | 0.41 | 0.33 | 0.52 | 1.09 | 2.38 | 6.59 | 26.75 |
| | High | 0.07 | 0.12 | 0.26 | 0.64 | 1.56 | 4.61 | 19.58 |
| | | | | Men | | | | |
| Argentina 2011 | Low | 0.12 | 0.26 | 0.52 | 0.88 | 2.07 | 5.93 | 21.20 |
| | High | 0.07 | 0.16 | 0.35 | 0.59 | 1.41 | 4.13 | 16.66 |
| Brazil 2011 | Low | 0.35 | 0.69 | 1.23 | 2.54 | 5.31 | 13.19 | 37.74 |
| | High | 0.18 | 0.40 | 0.91 | 2.21 | 5.43 | 15.75 | 52.19 |
| Chile 2018 | Low | 0.10 | 0.30 | 0.34 | 0.47 | 0.75 | 2.24 | 10.73 |
| | High | 0.05 | 0.08 | 0.16 | 0.25 | 0.49 | 1.20 | 7.47 |
| Colombia 2019 | Low | 0.47 | 0.59 | 0.74 | 1.25 | 3.11 | 7.40 | 23.93 |
| | High | 0.16 | 0.26 | 0.37 | 0.69 | 1.93 | 5.39 | 18.94 |
| Ecuador 2011 | Low | 0.30 | 0.38 | 0.32 | 0.50 | 0.91 | 2.83 | 13.21 |
| | High | 0.15 | 0.29 | 0.25 | 0.48 | 0.74 | 2.50 | 15.31 |
| Mexico 2011 | Low | 0.10 | 0.18 | 0.22 | 0.27 | 0.43 | 0.94 | 3.64 |
| | High | 0.05 | 0.09 | 0.14 | 0.18 | 0.33 | 0.85 | 3.48 |
| Peru 2018 | Low | 0.81 | 0.74 | 0.98 | 1.73 | 3.28 | 8.16 | 29.85 |
| | High | 0.14 | 0.24 | 0.39 | 0.84 | 2.18 | 5.98 | 25.91 |

*Note:* Authors' calculations based on census and death certificate data. See Appendix Table S1 for details on data sources.

Table S14: Mortality rates ratios by sex, age and education - Communicable

| Country | 20-29 | 30-39 | 40-49 | 50-59 | 60-69 | 70-79 | 80+ |
|---|---|---|---|---|---|---|---|
| Women | | | | | | | |
| Argentina 2011 | 2.16 | 2.19 | 1.90 | 1.74 | 1.54 | 1.43 | 1.25 |
| Brazil 2011 | 2.14 | 1.81 | 1.44 | 1.28 | 1.18 | 1.07 | 0.89 |
| Chile 2018 | 21.14 | 3.91 | 2.32 | 2.67 | 1.84 | 1.87 | 1.52 |
| Colombia 2019 | 3.94 | 2.97 | 2.25 | 2.21 | 1.93 | 1.54 | 1.43 |
| Ecuador 2011 | 1.63 | 1.90 | 1.53 | 1.35 | 1.59 | 1.35 | 1.15 |
| Mexico 2011 | 3.58 | 2.94 | 2.25 | 1.85 | 1.42 | 1.40 | 1.21 |
| Peru 2018 | 5.89 | 2.75 | 1.98 | 1.70 | 1.53 | 1.43 | 1.37 |
| Median | 3.58 | 2.75 | 1.98 | 1.74 | 1.54 | 1.43 | 1.25 |
| Max/Min | 13.00 | 2.16 | 1.61 | 2.08 | 1.64 | 1.75 | 1.72 |
| Men | | | | | | | |
| Argentina 2011 | 1.68 | 1.65 | 1.48 | 1.50 | 1.47 | 1.44 | 1.27 |
| Brazil 2011 | 1.89 | 1.71 | 1.35 | 1.15 | 0.98 | 0.84 | 0.72 |
| Chile 2018 | 2.23 | 3.69 | 2.17 | 1.88 | 1.54 | 1.86 | 1.44 |
| Colombia 2019 | 2.91 | 2.31 | 2.01 | 1.82 | 1.61 | 1.37 | 1.26 |
| Ecuador 2011 | 2.06 | 1.29 | 1.31 | 1.05 | 1.24 | 1.13 | 0.86 |
| Mexico 2011 | 2.06 | 1.87 | 1.54 | 1.50 | 1.30 | 1.10 | 1.05 |
| Peru 2018 | 5.74 | 3.08 | 2.50 | 2.05 | 1.50 | 1.36 | 1.15 |
| Median | 2.06 | 1.87 | 1.54 | 1.50 | 1.47 | 1.36 | 1.15 |
| Max/Min | 3.42 | 2.85 | 1.91 | 1.95 | 1.64 | 2.22 | 1.98 |

*Note:* Authors' calculations based on census and death certificate data. See Appendix Table S1 for details on data sources.

#### Maternal deaths

Figure S2: Maternal Deaths

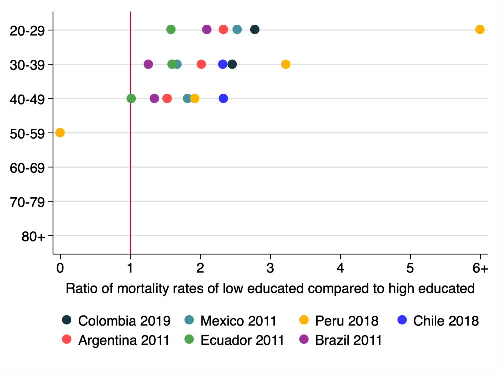

Women

*Note:* Authors' calculations based on census and death certificate data. See Appendix Table S1 for details on data sources.

Table S15: Mortality rates by sex, age and education - Maternal deaths

| Country | Education | 20-29 | 30-39 | 40-49 |
|---|---|---|---|---|
| Women | | | | |
| Argentina 2011 | Low | 0.05 | 0.07 | 0.02 |
| | High | 0.02 | 0.03 | 0.01 |
| Brazil 2011 | Low | 0.06 | 0.05 | 0.01 |
| | High | 0.03 | 0.04 | 0.01 |
| Chile 2018 | Low | 0.03 | 0.04 | 0.01 |
| | High | 0.01 | 0.02 | 0.00 |
| Colombia 2019 | Low | 0.12 | 0.09 | 0.02 |
| | High | 0.04 | 0.04 | 0.01 |
| Ecuador 2011 | Low | 0.09 | 0.12 | 0.03 |
| | High | 0.05 | 0.08 | 0.03 |
| Mexico 2011 | Low | 0.03 | 0.03 | 0.01 |
| | High | 0.01 | 0.02 | 0.00 |
| Peru 2018 | Low | 0.04 | 0.03 | 0.01 |
| | High | 0.01 | 0.01 | 0.00 |

*Note:* Authors' calculations based on census and death certificate data. See Appendix Table S1 for details on data sources.

Table S16: Mortality rates ratios by sex, age and education - Maternal deaths

| Country | 20-29 | 30-39 | 40-49 |
|---|---|---|---|
| Women | | | |
| Argentina 2011 | 2.33 | 2.02 | 1.53 |
| Brazil 2011 | 2.09 | 1.26 | 1.35 |
| Chile 2018 | 2.10 | 2.32 | 2.33 |
| Colombia 2019 | 2.78 | 2.46 | 1.53 |
| Ecuador 2011 | 1.58 | 1.60 | 1.01 |
| Mexico 2011 | 2.53 | 1.67 | 1.82 |
| Peru 2018 | 6.25 | 3.22 | 1.92 |
| Median | 2.33 | 2.02 | 1.53 |
| Max/Min | 3.95 | 2.56 | 2.30 |

*Note:* Authors' calculations based on census and death certificate data. See Appendix Table S1 for details on data sources.

Nutritional

Figure S3: Nutritional

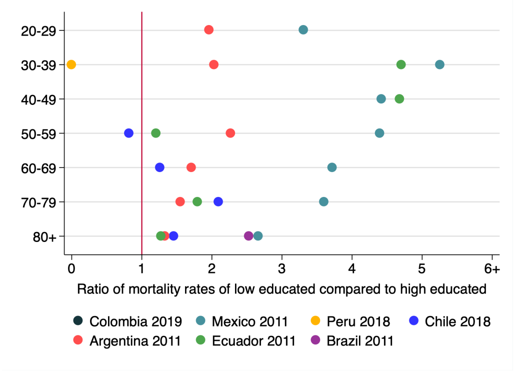

Women

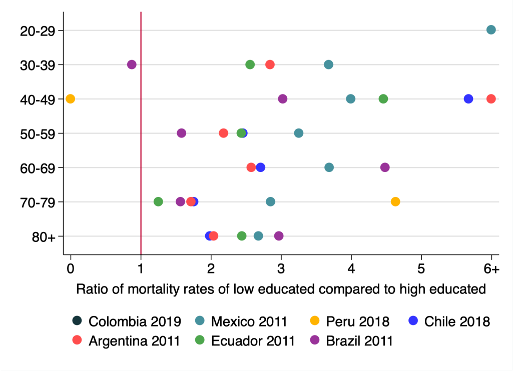

Men

*Note:* Authors' calculations based on census and death certificate data. See Appendix Table S1 for details on data sources.

Table S17: Mortality rates by sex, age and education - Nutritional causes

| Country | Education | 20-29 | 30-39 | 40-49 | 50-59 | 60-69 | 70-79 | 80+ |
|---|---|---|---|---|---|---|---|---|
| | | | | Women | | | | |
| Argentina 2011 | Low | 0.00 | 0.00 | 0.01 | 0.01 | 0.02 | 0.09 | 0.54 |
| | High | 0.00 | 0.00 | 0.00 | 0.00 | 0.01 | 0.06 | 0.41 |
| Brazil 2011 | Low | 0.00 | 0.00 | 0.00 | 0.00 | 0.00 | 0.01 | 0.07 |
| | High | 0.00 | 0.00 | 0.00 | 0.00 | 0.00 | 0.00 | 0.03 |
| Chile 2018 | Low | 0.01 | 0.00 | 0.00 | 0.00 | 0.02 | 0.09 | 0.67 |
| | High | 0.00 | 0.00 | 0.00 | 0.00 | 0.02 | 0.04 | 0.46 |
| Colombia 2019 | Low | 0.00 | 0.00 | 0.00 | 0.00 | 0.00 | 0.00 | 0.01 |
| | High | 0.00 | 0.00 | 0.00 | 0.00 | 0.00 | 0.00 | 0.00 |
| Ecuador 2011 | Low | 0.00 | 0.01 | 0.01 | 0.02 | 0.05 | 0.24 | 2.85 |
| | High | 0.00 | 0.00 | 0.00 | 0.02 | 0.00 | 0.13 | 2.23 |
| Mexico 2011 | Low | 0.00 | 0.01 | 0.01 | 0.02 | 0.05 | 0.20 | 1.73 |
| | High | 0.00 | 0.00 | 0.00 | 0.00 | 0.01 | 0.06 | 0.65 |
| Peru 2018 | Low | 0.00 | 0.00 | 0.00 | 0.00 | 0.00 | 0.02 | 0.09 |
| | High | 0.00 | 0.00 | 0.00 | 0.00 | 0.00 | 0.00 | 0.00 |
| | | | | Men | | | | |
| Argentina 2011 | Low | 0.00 | 0.00 | 0.01 | 0.01 | 0.05 | 0.19 | 0.86 |
| | High | 0.00 | 0.00 | 0.00 | 0.01 | 0.02 | 0.11 | 0.42 |
| Brazil 2011 | Low | 0.00 | 0.00 | 0.00 | 0.00 | 0.00 | 0.02 | 0.08 |
| | High | 0.00 | 0.00 | 0.00 | 0.00 | 0.00 | 0.01 | 0.03 |
| Chile 2018 | Low | 0.01 | 0.01 | 0.01 | 0.02 | 0.04 | 0.13 | 0.83 |
| | High | 0.00 | 0.00 | 0.00 | 0.01 | 0.01 | 0.08 | 0.42 |
| Colombia 2019 | Low | 0.00 | 0.00 | 0.00 | 0.00 | 0.00 | 0.00 | 0.00 |
| | High | 0.00 | 0.00 | 0.00 | 0.00 | 0.00 | 0.00 | 0.00 |
| Ecuador 2011 | Low | 0.01 | 0.01 | 0.01 | 0.03 | 0.05 | 0.41 | 2.35 |
| | High | 0.00 | 0.00 | 0.00 | 0.01 | 0.00 | 0.33 | 0.96 |
| Mexico 2011 | Low | 0.01 | 0.01 | 0.02 | 0.03 | 0.08 | 0.27 | 1.86 |
| | High | 0.00 | 0.00 | 0.00 | 0.01 | 0.02 | 0.09 | 0.70 |
| Peru 2018 | Low | 0.00 | 0.00 | 0.00 | 0.00 | 0.00 | 0.02 | 0.06 |
| | High | 0.00 | 0.00 | 0.00 | 0.00 | 0.00 | 0.00 | 0.00 |

*Note:* Authors' calculations based on census and death certificate data. See Appendix Table S1 for details on data sources.

Table S18: Mortality rates ratios by sex, age and education - Nutritional causes

| Country | 20-29 | 30-39 | 40-49 | 50-59 | 60-69 | 70-79 | 80+ |
|---|---|---|---|---|---|---|---|
| | | | Women | | | | |
| Argentina 2011 | 1.96 | 2.03 | - | 2.27 | 1.71 | 1.56 | 1.33 |
| Brazil 2011 | - | - | - | - | - | - | 2.53 |
| Chile 2018 | - | - | - | 0.82 | 1.26 | 2.09 | 1.46 |
| Colombia 2019 | - | - | - | - | - | - | - |
| Ecuador 2011 | - | 4.70 | 4.68 | 1.20 | - | 1.80 | 1.28 |
| Mexico 2011 | 3.31 | 5.25 | 4.42 | 4.40 | 3.72 | 3.60 | 2.66 |
| Peru 2018 | - | - | - | - | - | - | - |
| Median | 2.63 | 4.70 | 4.55 | 1.74 | 1.71 | 1.94 | 1.46 |
| Max/Min | 1.69 | 2.59 | 1.06 | 5.38 | 2.95 | 2.31 | 2.08 |
| Men | | | | | | | |
| Argentina 2011 | - | 2.84 | 6.07 | 2.18 | 2.58 | 1.72 | 2.04 |
| Brazil 2011 | - | 0.87 | 3.02 | 1.58 | 4.49 | 1.57 | 2.97 |
| Chile 2018 | - | - | 5.67 | 2.45 | 2.71 | 1.75 | 1.98 |
| Colombia 2019 | - | - | - | - | - | - | - |
| Ecuador 2011 | - | 2.56 | 4.46 | 2.43 | - | 1.25 | 2.44 |
| Mexico 2011 | 13.80 | 3.68 | 4.00 | 3.25 | 3.69 | 2.85 | 2.68 |
| Peru 2018 | - | - | - | - | - | 4.63 | - |
| Median | 13.80 | 2.70 | 4.46 | 2.43 | 3.20 | 1.74 | 2.44 |
| Max/Min | 1.00 | 4.22 | 2.01 | 2.06 | 1.74 | 3.70 | 1.50 |

*Note:* Authors' calculations based on census and death certificate data. See Appendix Table S1 for details on data sources.

#### Neoplasms

Figure S4: Neoplasms

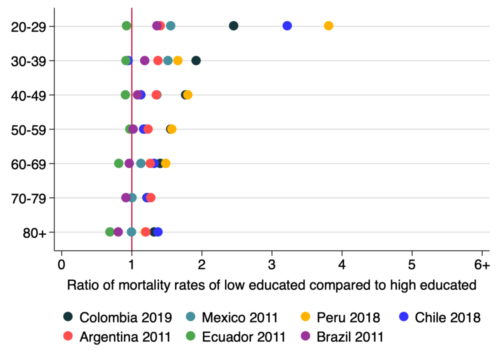

Women

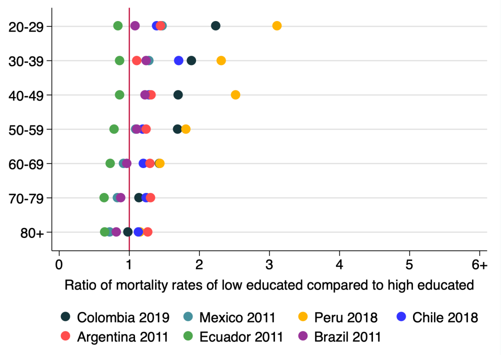

Men

*Note:* Authors' calculations based on census and death certificate data. See Appendix Table S1 for details on data sources.

Table S19: Mortality rates by sex, age and education - Neoplasms

| Country | Education | 20-29 | 30-39 | 40-49 | 50-59 | 60-69 | 70-79 | 80+ |
|---|---|---|---|---|---|---|---|---|
| | | | | Women | | | | |
| Argentina 2011 | Low | 0.10 | 0.35 | 0.96 | 2.29 | 4.22 | 6.89 | 12.05 |
| | High | 0.07 | 0.26 | 0.71 | 1.85 | 3.33 | 5.39 | 9.98 |
| Brazil 2011 | Low | 0.07 | 0.22 | 0.62 | 1.38 | 2.44 | 4.16 | 6.89 |
| | High | 0.05 | 0.19 | 0.57 | 1.35 | 2.52 | 4.51 | 8.53 |
| Chile 2018 | Low | 0.18 | 0.20 | 0.64 | 1.68 | 4.05 | 7.47 | 14.87 |
| | High | 0.06 | 0.21 | 0.57 | 1.43 | 3.06 | 6.07 | 10.76 |
| Colombia 2019 | Low | 0.17 | 0.43 | 0.93 | 1.67 | 3.20 | 5.83 | 10.68 |
| | High | 0.07 | 0.22 | 0.53 | 1.07 | 2.27 | 4.76 | 8.07 |
| Ecuador 2011 | Low | 0.07 | 0.25 | 0.65 | 1.49 | 2.84 | 6.99 | 15.55 |
| | High | 0.07 | 0.27 | 0.70 | 1.53 | 3.48 | 7.60 | 22.56 |
| Mexico 2011 | Low | 0.05 | 0.14 | 0.37 | 0.79 | 1.45 | 2.47 | 3.98 |
| | High | 0.03 | 0.09 | 0.27 | 0.66 | 1.28 | 2.46 | 3.99 |
| Peru 2018 | Low | 0.11 | 0.16 | 0.44 | 0.82 | 1.46 | 2.48 | 4.20 |
| | High | 0.03 | 0.09 | 0.24 | 0.52 | 0.98 | 1.98 | 3.51 |
| | | | | Men | | | | |
| Argentina 2011 | Low | 0.13 | 0.21 | 0.75 | 2.56 | 6.72 | 13.12 | 22.31 |
| | High | 0.09 | 0.19 | 0.57 | 2.06 | 5.17 | 10.06 | 17.63 |
| Brazil 2011 | Low | 0.07 | 0.14 | 0.53 | 1.66 | 3.66 | 6.97 | 12.39 |
| | High | 0.06 | 0.11 | 0.43 | 1.49 | 3.76 | 7.89 | 15.16 |
| Chile 2018 | Low | 0.12 | 0.25 | 0.53 | 1.70 | 5.28 | 12.62 | 24.85 |
| | High | 0.08 | 0.15 | 0.41 | 1.41 | 4.38 | 10.13 | 21.87 |
| Colombia 2019 | Low | 0.16 | 0.25 | 0.54 | 1.43 | 3.80 | 8.20 | 15.50 |
| | High | 0.07 | 0.13 | 0.32 | 0.84 | 2.66 | 7.16 | 15.79 |
| Ecuador 2011 | Low | 0.10 | 0.14 | 0.36 | 0.93 | 2.65 | 7.59 | 19.73 |
| | High | 0.12 | 0.16 | 0.41 | 1.18 | 3.63 | 11.72 | 30.15 |
| Mexico 2011 | Low | 0.07 | 0.09 | 0.22 | 0.60 | 1.51 | 3.31 | 6.01 |
| | High | 0.05 | 0.07 | 0.17 | 0.55 | 1.64 | 3.97 | 8.32 |
| Peru 2018 | Low | 0.11 | 0.15 | 0.32 | 0.62 | 1.23 | 2.89 | 5.42 |
| | High | 0.03 | 0.07 | 0.12 | 0.34 | 0.85 | 2.31 | 4.68 |

*Note:* Authors' calculations based on census and death certificate data. See Appendix Table S1 for details on data sources.

Table S20: Mortality rates ratios by sex, age and education - Neoplasms

| Country | 20-29 | 30-39 | 40-49 | 50-59 | 60-69 | 70-79 | 80+ |
|---|---|---|---|---|---|---|---|
| | | | Women | | | | |
| Argentina 2011 | 1.41 | 1.38 | 1.36 | 1.24 | 1.27 | 1.28 | 1.21 |
| Brazil 2011 | 1.36 | 1.19 | 1.09 | 1.03 | 0.97 | 0.92 | 0.81 |
| Chile 2018 | 3.22 | 0.95 | 1.13 | 1.18 | 1.32 | 1.23 | 1.38 |
| Colombia 2019 | 2.46 | 1.93 | 1.77 | 1.56 | 1.41 | 1.22 | 1.32 |
| Ecuador 2011 | 0.93 | 0.92 | 0.92 | 0.97 | 0.82 | 0.92 | 0.69 |
| Mexico 2011 | 1.56 | 1.52 | 1.36 | 1.20 | 1.13 | 1.00 | 1.00 |
| Peru 2018 | 3.81 | 1.66 | 1.81 | 1.57 | 1.49 | 1.25 | 1.19 |
| Median | 1.56 | 1.38 | 1.36 | 1.20 | 1.27 | 1.22 | 1.19 |
| Max/Min | 4.10 | 2.10 | 1.97 | 1.62 | 1.82 | 1.39 | 2.00 |
| Men | | | | | | | |
| Argentina 2011 | 1.45 | 1.11 | 1.32 | 1.24 | 1.30 | 1.30 | 1.27 |
| Brazil 2011 | 1.09 | 1.25 | 1.23 | 1.11 | 0.97 | 0.88 | 0.82 |
| Chile 2018 | 1.40 | 1.71 | 1.29 | 1.20 | 1.21 | 1.25 | 1.14 |
| Colombia 2019 | 2.24 | 1.89 | 1.70 | 1.69 | 1.43 | 1.14 | 0.98 |
| Ecuador 2011 | 0.84 | 0.86 | 0.87 | 0.78 | 0.73 | 0.65 | 0.65 |
| Mexico 2011 | 1.47 | 1.28 | 1.28 | 1.09 | 0.92 | 0.84 | 0.72 |
| Peru 2018 | 3.11 | 2.32 | 2.52 | 1.81 | 1.44 | 1.25 | 1.16 |
| Median | 1.45 | 1.28 | 1.29 | 1.20 | 1.21 | 1.14 | 0.98 |
| Max/Min | 3.71 | 2.68 | 2.90 | 2.31 | 1.98 | 2.01 | 1.93 |

*Note:* Authors' calculations based on census and death certificate data. See Appendix Table S1 for details on data sources.

#### Cardiovascular diseases

Figure S5: Cardiovascular diseases

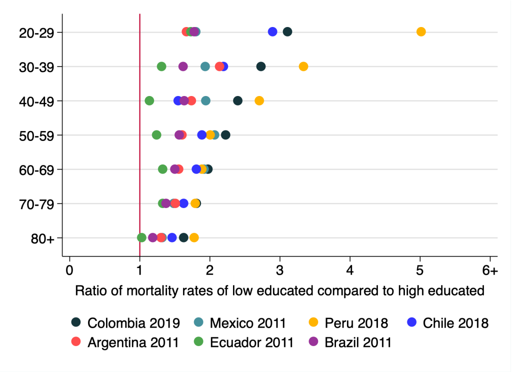

Women

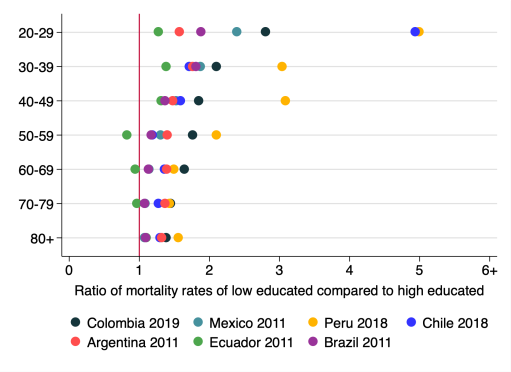

Men

*Note:* Authors' calculations based on census and death certificate data. See Appendix Table S1 for details on data sources.

Table S21: Mortality rates by sex, age and education - Cardiovascular diseases

| Country | Education | 20-29 | 30-39 | 40-49 | 50-59 | 60-69 | 70-79 | 80+ |
|---|---|---|---|---|---|---|---|---|
| | | | | Women | | | | |
| Argentina 2011 | Low | 0.06 | 0.15 | 0.47 | 1.24 | 3.42 | 10.37 | 48.30 |
| | High | 0.04 | 0.07 | 0.27 | 0.77 | 2.19 | 6.85 | 36.93 |
| Brazil 2011 | Low | 0.11 | 0.27 | 0.87 | 2.14 | 5.07 | 12.45 | 37.64 |
| | High | 0.06 | 0.16 | 0.53 | 1.37 | 3.37 | 9.05 | 31.67 |
| Chile 2018 | Low | 0.06 | 0.11 | 0.28 | 0.77 | 2.21 | 6.53 | 31.94 |
| | High | 0.02 | 0.05 | 0.18 | 0.41 | 1.22 | 4.00 | 21.80 |
| Colombia 2019 | Low | 0.23 | 0.37 | 0.85 | 1.85 | 4.82 | 13.11 | 53.31 |
| | High | 0.07 | 0.14 | 0.35 | 0.83 | 2.44 | 7.23 | 32.73 |
| Ecuador 2011 | Low | 0.08 | 0.14 | 0.36 | 1.03 | 2.66 | 8.15 | 44.45 |
| | High | 0.05 | 0.10 | 0.32 | 0.83 | 2.00 | 6.12 | 43.00 |
| Mexico 2011 | Low | 0.03 | 0.06 | 0.19 | 0.59 | 1.64 | 4.93 | 20.91 |
| | High | 0.02 | 0.03 | 0.10 | 0.29 | 0.86 | 3.31 | 15.78 |
| Peru 2018 | Low | 0.18 | 0.17 | 0.37 | 0.66 | 1.48 | 3.55 | 13.51 |
| | High | 0.04 | 0.05 | 0.14 | 0.33 | 0.78 | 1.98 | 7.60 |
| | | | | Men | | | | |
| Argentina 2011 | Low | 0.09 | 0.27 | 0.94 | 2.96 | 7.60 | 18.29 | 55.05 |
| | High | 0.06 | 0.15 | 0.63 | 2.11 | 5.45 | 13.31 | 41.53 |
| Brazil 2011 | Low | 0.18 | 0.47 | 1.33 | 3.47 | 7.87 | 17.20 | 42.31 |
| | High | 0.09 | 0.26 | 0.97 | 2.97 | 6.95 | 15.95 | 38.57 |
| Chile 2018 | Low | 0.13 | 0.22 | 0.67 | 1.52 | 4.28 | 10.65 | 35.44 |
| | High | 0.03 | 0.13 | 0.42 | 1.28 | 3.15 | 8.33 | 27.28 |
| Colombia 2019 | Low | 0.28 | 0.46 | 0.97 | 2.68 | 7.79 | 19.50 | 61.62 |
| | High | 0.10 | 0.22 | 0.52 | 1.52 | 4.74 | 13.45 | 44.44 |
| Ecuador 2011 | Low | 0.13 | 0.25 | 0.60 | 1.36 | 3.62 | 11.12 | 42.16 |
| | High | 0.10 | 0.18 | 0.46 | 1.64 | 3.83 | 11.51 | 38.35 |
| Mexico 2011 | Low | 0.07 | 0.15 | 0.40 | 1.01 | 2.47 | 6.14 | 19.98 |
| | High | 0.03 | 0.08 | 0.26 | 0.77 | 2.16 | 5.65 | 18.58 |
| Peru 2018 | Low | 0.35 | 0.31 | 0.59 | 1.06 | 1.89 | 4.65 | 14.96 |
| | High | 0.07 | 0.10 | 0.19 | 0.50 | 1.26 | 3.24 | 9.58 |

*Note:* Authors' calculations based on census and death certificate data. See Appendix Table S1 for details on data sources.

Table S22: Mortality rates ratios by sex, age and education - Cardiovascular diseases

| Country | 20-29 | 30-39 | 40-49 | 50-59 | 60-69 | 70-79 | 80+ |
|---|---|---|---|---|---|---|---|
| Women | | | | | | | |
| Argentina 2011 | 1.67 | 2.14 | 1.74 | 1.61 | 1.56 | 1.51 | 1.31 |
| Brazil 2011 | 1.78 | 1.62 | 1.64 | 1.57 | 1.50 | 1.38 | 1.19 |
| Chile 2018 | 2.90 | 2.20 | 1.55 | 1.89 | 1.81 | 1.63 | 1.46 |
| Colombia 2019 | 3.11 | 2.73 | 2.41 | 2.23 | 1.97 | 1.81 | 1.63 |
| Ecuador 2011 | 1.73 | 1.32 | 1.14 | 1.24 | 1.33 | 1.33 | 1.03 |
| Mexico 2011 | 1.81 | 1.94 | 1.94 | 2.07 | 1.92 | 1.49 | 1.33 |
| Peru 2018 | 5.02 | 3.34 | 2.71 | 2.01 | 1.89 | 1.79 | 1.78 |
| Median | 1.81 | 2.14 | 1.74 | 1.89 | 1.81 | 1.51 | 1.33 |
| Max/Min | 3.00 | 2.54 | 2.37 | 1.79 | 1.49 | 1.36 | 1.72 |
| Men | | | | | | | |
| Argentina 2011 | 1.57 | 1.76 | 1.48 | 1.40 | 1.40 | 1.37 | 1.33 |
| Brazil 2011 | 1.88 | 1.81 | 1.37 | 1.17 | 1.13 | 1.08 | 1.10 |
| Chile 2018 | 4.94 | 1.71 | 1.59 | 1.19 | 1.36 | 1.28 | 1.30 |
| Colombia 2019 | 2.81 | 2.11 | 1.85 | 1.77 | 1.64 | 1.45 | 1.39 |
| Ecuador 2011 | 1.28 | 1.39 | 1.31 | 0.83 | 0.94 | 0.97 | 1.10 |
| Mexico 2011 | 2.39 | 1.87 | 1.53 | 1.31 | 1.14 | 1.09 | 1.08 |
| Peru 2018 | 4.99 | 3.04 | 3.09 | 2.10 | 1.50 | 1.44 | 1.56 |
| Median | 2.39 | 1.81 | 1.53 | 1.31 | 1.36 | 1.28 | 1.30 |
| Max/Min | 3.90 | 2.19 | 2.35 | 2.55 | 1.74 | 1.50 | 1.45 |

*Note:* Authors' calculations based on census and death certificate data. See Apperndix Table S1 for details on data sources.

#### Diabetes

Figure S6: Diabetes

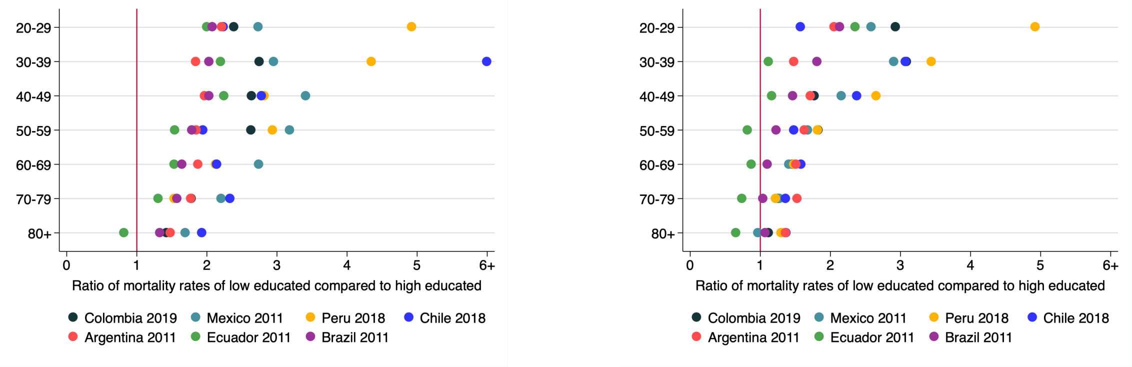

Women Men

*Note:* Authors' calculations based on census and death certificate data. See Appendix Table S1 for details on data sources.

Table S23: Mortality rates by sex, age and education - Diabetes

| Country | Education | 20-29 | 30-39 | 40-49 | 50-59 | 60-69 | 70-79 | 80+ |
|---|---|---|---|---|---|---|---|---|
| | | | | Women | | | | |
| Argentina 2011 | Low | 0.02 | 0.04 | 0.10 | 0.32 | 0.95 | 2.08 | 5.19 |
| | High | 0.01 | 0.02 | 0.05 | 0.17 | 0.51 | 1.18 | 3.51 |
| Brazil 2011 | Low | 0.03 | 0.07 | 0.18 | 0.48 | 1.26 | 2.95 | 6.87 |
| | High | 0.01 | 0.03 | 0.09 | 0.27 | 0.77 | 1.87 | 5.16 |
| Chile 2018 | Low | 0.01 | 0.03 | 0.06 | 0.18 | 0.55 | 1.53 | 4.79 |
| | High | 0.00 | 0.00 | 0.02 | 0.09 | 0.26 | 0.66 | 2.48 |
| Colombia 2019 | Low | 0.05 | 0.10 | 0.22 | 0.53 | 1.56 | 3.64 | 10.69 |
| | High | 0.02 | 0.04 | 0.08 | 0.20 | 0.73 | 2.04 | 7.48 |
| Ecuador 2011 | Low | 0.04 | 0.08 | 0.28 | 0.84 | 2.16 | 5.54 | 14.86 |
| | High | 0.02 | 0.04 | 0.12 | 0.54 | 1.40 | 4.25 | 18.21 |
| Mexico 2011 | Low | 0.03 | 0.06 | 0.24 | 0.89 | 2.23 | 4.04 | 7.13 |
| | High | 0.01 | 0.02 | 0.07 | 0.28 | 0.81 | 1.83 | 4.21 |
| Peru 2018 | Low | 0.05 | 0.07 | 0.12 | 0.27 | 0.59 | 0.96 | 2.54 |
| | High | 0.01 | 0.02 | 0.04 | 0.09 | 0.28 | 0.63 | 1.32 |
| | | | | Men | | | | |
| Argentina 2011 | Low | 0.02 | 0.03 | 0.15 | 0.52 | 1.41 | 3.34 | 7.88 |
| | High | 0.01 | 0.02 | 0.09 | 0.32 | 0.93 | 2.18 | 5.78 |
| Brazil 2011 | Low | 0.03 | 0.07 | 0.21 | 0.61 | 1.52 | 3.41 | 7.91 |
| | High | 0.02 | 0.04 | 0.14 | 0.50 | 1.38 | 3.27 | 7.38 |
| Chile 2018 | Low | 0.01 | 0.03 | 0.09 | 0.24 | 0.75 | 2.06 | 5.97 |
| | High | 0.00 | 0.01 | 0.04 | 0.16 | 0.48 | 1.51 | 4.37 |
| Colombia 2019 | Low | 0.05 | 0.09 | 0.19 | 0.68 | 1.92 | 4.86 | 13.62 |
| | High | 0.02 | 0.03 | 0.11 | 0.37 | 1.31 | 3.85 | 12.19 |
| Ecuador 2011 | Low | 0.05 | 0.07 | 0.22 | 0.74 | 1.91 | 4.19 | 12.24 |
| | High | 0.02 | 0.06 | 0.19 | 0.91 | 2.18 | 5.65 | 18.81 |
| Mexico 2011 | Low | 0.05 | 0.10 | 0.36 | 1.05 | 2.29 | 4.04 | 6.65 |
| | High | 0.02 | 0.03 | 0.17 | 0.63 | 1.63 | 3.17 | 6.87 |
| Peru 2018 | Low | 0.04 | 0.07 | 0.12 | 0.31 | 0.61 | 1.19 | 3.07 |
| | High | 0.01 | 0.02 | 0.05 | 0.17 | 0.41 | 0.97 | 2.36 |

*Note:* Authors' calculations based on census and death certificate data. See Appendix Table S1 for details on data sources.

Table S24: Mortality rates ratios by sex, age and education - Diabetes

| Country | 20-29 | 30-39 | 40-49 | 50-59 | 60-69 | 70-79 | 80+ |
|---|---|---|---|---|---|---|---|
| | | | Women | | | | |
| Argentina 2011 | 2.21 | 1.84 | 1.97 | 1.85 | 1.88 | 1.77 | 1.48 |
| Brazil 2011 | 2.08 | 2.03 | 2.03 | 1.79 | 1.65 | 1.58 | 1.33 |
| Chile 2018 | 2.24 | 6.59 | 2.78 | 1.95 | 2.15 | 2.33 | 1.93 |
| Colombia 2019 | 2.39 | 2.75 | 2.64 | 2.63 | 2.14 | 1.78 | 1.43 |
| Ecuador 2011 | 2.00 | 2.20 | 2.25 | 1.54 | 1.54 | 1.31 | 0.82 |
| Mexico 2011 | 2.74 | 2.95 | 3.41 | 3.19 | 2.74 | 2.21 | 1.70 |
| Peru 2018 | 4.93 | 4.35 | 2.82 | 2.94 | 2.13 | 1.53 | 1.93 |
| Median | 2.24 | 2.75 | 2.64 | 1.95 | 2.13 | 1.77 | 1.48 |
| Max/Min | 2.46 | 3.57 | 1.73 | 2.06 | 1.79 | 1.79 | 2.37 |
| Men | | | | | | | |
| Argentina 2011 | 2.05 | 1.48 | 1.72 | 1.63 | 1.51 | 1.53 | 1.36 |
| Brazil 2011 | 2.14 | 1.81 | 1.47 | 1.23 | 1.10 | 1.04 | 1.07 |
| Chile 2018 | 1.57 | 3.07 | 2.38 | 1.48 | 1.58 | 1.36 | 1.37 |
| Colombia 2019 | 2.93 | 3.09 | 1.77 | 1.83 | 1.46 | 1.26 | 1.12 |
| Ecuador 2011 | 2.36 | 1.12 | 1.17 | 0.82 | 0.88 | 0.74 | 0.65 |
| Mexico 2011 | 2.58 | 2.91 | 2.16 | 1.68 | 1.41 | 1.28 | 0.97 |
| Peru 2018 | 4.92 | 3.45 | 2.65 | 1.82 | 1.48 | 1.22 | 1.30 |
| Median | 2.36 | 2.91 | 1.77 | 1.63 | 1.46 | 1.26 | 1.12 |
| Max/Min | 3.13 | 3.09 | 2.28 | 2.23 | 1.80 | 2.07 | 2.10 |

*Note:* Authors' calculations based on census and death certificate data. See Appendix Table S1 for details on data sources.

Unintentional injuries

Table S25: Mortality rates by sex, age and education - Unintentional injuries

| Country | Education | 20-29 | 30-39 | 40-49 | 50-59 | 60-69 | 70-79 | 80+ |
|---|---|---|---|---|---|---|---|---|
| | | | | Women | | | | |
| Argentina 2011 | Low | 0.16 | 0.15 | 0.15 | 0.21 | 0.27 | 0.50 | 1.74 |
| | High | 0.11 | 0.10 | 0.11 | 0.16 | 0.19 | 0.37 | 1.32 |
| Brazil 2011 | Low | 0.13 | 0.11 | 0.12 | 0.13 | 0.18 | 0.35 | 0.94 |
| | High | 0.11 | 0.09 | 0.10 | 0.13 | 0.20 | 0.33 | 1.04 |
| Chile 2018 | Low | 0.13 | 0.10 | 0.12 | 0.13 | 0.19 | 0.45 | 2.40 |
| | High | 0.08 | 0.06 | 0.07 | 0.09 | 0.15 | 0.39 | 1.71 |
| Colombia 2019 | Low | 0.06 | 0.07 | 0.06 | 0.07 | 0.08 | 0.16 | 0.39 |
| | High | 0.07 | 0.06 | 0.05 | 0.06 | 0.07 | 0.18 | 0.35 |
| Ecuador 2011 | Low | 0.15 | 0.15 | 0.19 | 0.26 | 0.39 | 0.82 | 2.69 |
| | High | 0.12 | 0.12 | 0.12 | 0.16 | 0.25 | 0.58 | 2.35 |
| Mexico 2011 | Low | 0.06 | 0.06 | 0.06 | 0.08 | 0.13 | 0.29 | 1.09 |
| | High | 0.05 | 0.04 | 0.05 | 0.06 | 0.08 | 0.19 | 0.62 |
| Peru 2018 | Low | 0.07 | 0.07 | 0.07 | 0.08 | 0.07 | 0.13 | 0.28 |
| | High | 0.02 | 0.02 | 0.02 | 0.02 | 0.02 | 0.06 | 0.15 |
| | | | | Men | | | | |
| Argentina 2011 | Low | 0.94 | 0.71 | 0.63 | 0.73 | 0.86 | 1.24 | 2.16 |
| | High | 0.56 | 0.45 | 0.41 | 0.53 | 0.68 | 0.84 | 1.51 |
| Brazil 2011 | Low | 0.88 | 0.82 | 0.80 | 0.81 | 0.79 | 0.94 | 1.47 |
| | High | 0.63 | 0.58 | 0.60 | 0.65 | 0.72 | 0.90 | 1.71 |
| Chile 2018 | Low | 0.72 | 0.77 | 0.65 | 0.85 | 0.85 | 1.21 | 2.64 |
| | High | 0.37 | 0.38 | 0.35 | 0.39 | 0.51 | 0.89 | 2.16 |
| Colombia 2019 | Low | 0.53 | 0.46 | 0.41 | 0.44 | 0.52 | 0.65 | 0.91 |
| | High | 0.38 | 0.30 | 0.28 | 0.27 | 0.35 | 0.54 | 0.86 |
| Ecuador 2011 | Low | 1.03 | 1.00 | 1.10 | 1.14 | 1.30 | 2.34 | 3.91 |
| | High | 1.02 | 0.79 | 0.69 | 0.75 | 0.81 | 1.97 | 3.86 |
| Mexico 2011 | Low | 0.47 | 0.44 | 0.43 | 0.47 | 0.53 | 0.68 | 1.40 |
| | High | 0.25 | 0.24 | 0.23 | 0.27 | 0.34 | 0.45 | 0.98 |
| Peru 2018 | Low | 0.50 | 0.36 | 0.34 | 0.36 | 0.26 | 0.34 | 0.47 |
| | High | 0.11 | 0.10 | 0.12 | 0.10 | 0.12 | 0.11 | 0.22 |

*Note:* Authors' calculations based on census and death certificate data. See Appendix Table S1 for details on data sources.

Table S26: Mortality rates ratios by sex, age and education - Unintentional injuries

| Country | 20-29 | 30-39 | 40-49 | 50-59 | 60-69 | 70-79 | 80+ |
|---|---|---|---|---|---|---|---|
| | | | Women | | | | |
| Argentina 2011 | 1.42 | 1.45 | 1.30 | 1.26 | 1.38 | 1.34 | 1.32 |
| Brazil 2011 | 1.14 | 1.19 | 1.18 | 1.04 | 0.93 | 1.07 | 0.90 |
| Chile 2018 | 1.66 | 1.57 | 1.75 | 1.55 | 1.31 | 1.17 | 1.40 |
| Colombia 2019 | 0.90 | 1.29 | 1.27 | 1.18 | 1.28 | 0.91 | 1.11 |
| Ecuador 2011 | 1.20 | 1.25 | 1.50 | 1.60 | 1.55 | 1.40 | 1.14 |
| Mexico 2011 | 1.12 | 1.25 | 1.32 | 1.30 | 1.69 | 1.50 | 1.77 |
| Peru 2018 | 2.79 | 3.73 | 3.24 | 4.09 | 2.90 | 2.09 | 1.85 |
| Median | 1.20 | 1.29 | 1.32 | 1.30 | 1.38 | 1.34 | 1.32 |
| Max/Min | 3.10 | 3.14 | 2.74 | 3.92 | 3.11 | 2.30 | 2.05 |
| Men | | | | | | | |
| Argentina 2011 | 1.67 | 1.56 | 1.55 | 1.37 | 1.25 | 1.47 | 1.42 |
| Brazil 2011 | 1.41 | 1.41 | 1.33 | 1.26 | 1.10 | 1.04 | 0.86 |
| Chile 2018 | 1.97 | 2.01 | 1.88 | 2.16 | 1.65 | 1.36 | 1.22 |
| Colombia 2019 | 1.41 | 1.54 | 1.48 | 1.63 | 1.50 | 1.20 | 1.06 |
| Ecuador 2011 | 1.01 | 1.28 | 1.60 | 1.51 | 1.61 | 1.19 | 1.01 |
| Mexico 2011 | 1.84 | 1.80 | 1.87 | 1.77 | 1.58 | 1.51 | 1.43 |
| Peru 2018 | 4.67 | 3.65 | 2.90 | 3.48 | 2.12 | 3.09 | 2.11 |
| Median | 1.67 | 1.56 | 1.60 | 1.63 | 1.58 | 1.36 | 1.22 |
| Max/Min | 4.64 | 2.86 | 2.17 | 2.77 | 1.94 | 2.97 | 2.45 |

*Note:* Authors' calculations based on census and death certificate data. See Appendix Table S1 for details on data sources.

Intentional injuries

Table S27: Mortality rates by sex, age and education - Intentional injuries

| Country | Education | 20-29 | 30-39 | 40-49 | 50-59 | 60-69 | 70-79 | 80+ |
|---|---|---|---|---|---|---|---|---|
| | | | | Women | | | | |
| Argentina 2011 | Low | 0.08 | 0.06 | 0.06 | 0.05 | 0.04 | 0.04 | 0.05 |
| | High | 0.04 | 0.04 | 0.04 | 0.05 | 0.03 | 0.03 | 0.04 |
| Brazil 2011 | Low | 0.16 | 0.12 | 0.09 | 0.07 | 0.05 | 0.06 | 0.06 |
| | High | 0.07 | 0.07 | 0.07 | 0.06 | 0.05 | 0.05 | 0.06 |
| Chile 2018 | Low | 0.15 | 0.09 | 0.06 | 0.05 | 0.03 | 0.02 | 0.02 |
| | High | 0.05 | 0.06 | 0.06 | 0.05 | 0.04 | 0.03 | 0.04 |
| Colombia 2019 | Low | 0.22 | 0.15 | 0.10 | 0.06 | 0.05 | 0.05 | 0.06 |
| | High | 0.10 | 0.08 | 0.06 | 0.04 | 0.05 | 0.05 | 0.03 |
| Ecuador 2011 | Low | 0.14 | 0.09 | 0.07 | 0.07 | 0.06 | 0.06 | 0.15 |
| | High | 0.09 | 0.06 | 0.06 | 0.05 | 0.07 | 0.04 | 0.12 |
| Mexico 2011 | Low | 0.07 | 0.05 | 0.04 | 0.03 | 0.02 | 0.02 | 0.04 |
| | High | 0.04 | 0.03 | 0.03 | 0.03 | 0.04 | 0.02 | 0.03 |
| Peru 2018 | Low | 0.02 | 0.00 | 0.01 | 0.00 | 0.00 | 0.01 | 0.00 |
| | High | 0.00 | 0.00 | 0.00 | 0.00 | 0.00 | 0.00 | 0.00 |
| | | | | Men | | | | |
| Argentina 2011 | Low | 0.49 | 0.31 | 0.21 | 0.21 | 0.24 | 0.32 | 0.45 |
| | High | 0.25 | 0.17 | 0.14 | 0.14 | 0.16 | 0.21 | 0.30 |
| Brazil 2011 | Low | 1.92 | 1.11 | 0.66 | 0.44 | 0.34 | 0.31 | 0.31 |
| | High | 0.71 | 0.58 | 0.45 | 0.35 | 0.35 | 0.28 | 0.41 |
| Chile 2018 | Low | 0.69 | 0.69 | 0.43 | 0.35 | 0.34 | 0.31 | 0.35 |
| | High | 0.25 | 0.27 | 0.25 | 0.21 | 0.16 | 0.23 | 0.24 |
| Colombia 2019 | Low | 2.52 | 1.73 | 0.98 | 0.61 | 0.45 | 0.30 | 0.31 |
| | High | 0.84 | 0.70 | 0.54 | 0.42 | 0.32 | 0.27 | 0.21 |
| Ecuador 2011 | Low | 0.87 | 0.70 | 0.50 | 0.43 | 0.33 | 0.33 | 0.41 |
| | High | 0.64 | 0.58 | 0.50 | 0.36 | 0.21 | 0.12 | 0.36 |
| Mexico 2011 | Low | 0.68 | 0.56 | 0.40 | 0.26 | 0.19 | 0.17 | 0.18 |
| | High | 0.27 | 0.32 | 0.26 | 0.21 | 0.20 | 0.16 | 0.21 |
| Peru 2018 | Low | 0.07 | 0.03 | 0.02 | 0.03 | 0.02 | 0.00 | 0.01 |
| | High | 0.01 | 0.01 | 0.01 | 0.01 | 0.00 | 0.00 | 0.00 |

*Note:* Authors' calculations based on census and death certificate data. See Appendix Table S1 for details on data sources.

Table S28: Mortality rates ratios by sex, age and education - Intentional injuries

| Country | 20-29 | 30-39 | 40-49 | 50-59 | 60-69 | 70-79 | 80+ |
|---|---|---|---|---|---|---|---|
| **Women** | | | | | | | |
| Argentina 2011 | 2.06 | 1.69 | 1.49 | 0.94 | 1.14 | 1.32 | 1.26 |
| Brazil 2011 | 2.11 | 1.66 | 1.19 | 1.07 | 1.09 | 1.19 | 1.06 |
| Chile 2018 | 2.92 | 1.54 | 0.93 | 0.90 | 0.88 | 0.61 | 0.52 |
| Colombia 2019 | 2.24 | 1.82 | 1.69 | 1.41 | 1.02 | 1.03 | 1.71 |
| Ecuador 2011 | 1.47 | 1.60 | 1.21 | 1.61 | 0.86 | 1.39 | 1.27 |
| Mexico 2011 | 1.58 | 1.44 | 1.25 | 0.79 | 0.68 | 1.02 | 1.12 |
| Peru 2018 | 5.28 | - | 4.61 | - | - | - | - |
| Median | 2.11 | 1.63 | 1.25 | 1.01 | 0.95 | 1.11 | 1.19 |
| Max/Min | 3.60 | 1.26 | 4.97 | 2.05 | 1.67 | 2.29 | 3.31 |
| **Men** | | | | | | | |
| Argentina 2011 | 1.96 | 1.87 | 1.49 | 1.54 | 1.45 | 1.52 | 1.51 |
| Brazil 2011 | 2.70 | 1.93 | 1.47 | 1.28 | 0.99 | 1.14 | 0.77 |
| Chile 2018 | 2.74 | 2.56 | 1.76 | 1.64 | 2.05 | 1.33 | 1.47 |
| Colombia 2019 | 2.98 | 2.47 | 1.80 | 1.44 | 1.41 | 1.09 | 1.45 |
| Ecuador 2011 | 1.37 | 1.20 | 0.99 | 1.19 | 1.63 | 2.71 | 1.15 |
| Mexico 2011 | 2.52 | 1.74 | 1.53 | 1.23 | 0.98 | 1.06 | 0.89 |
| Peru 2018 | 6.54 | 3.04 | 2.90 | 4.05 | 9.63 | 0.77 | - |
| Median | 2.70 | 1.93 | 1.53 | 1.44 | 1.45 | 1.14 | 1.30 |
| Max/Min | 4.79 | 2.52 | 2.92 | 3.40 | 9.80 | 3.51 | 1.96 |

*Note:* Authors' calculations based on census and death certificate data. See Appendix Table S1 for details on data sources.

#### Suicides

Figure S7: Suicides

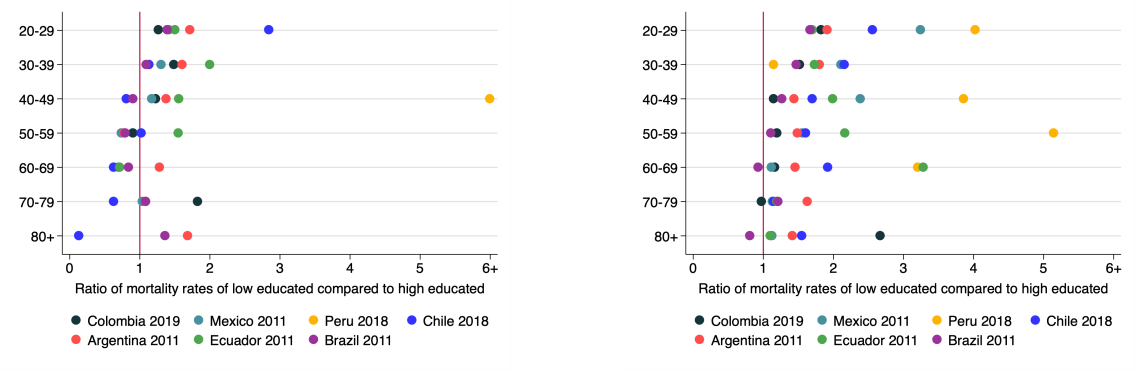

Women

Men

*Note:* Authors' calculations based on census and death certificate data. See Appendix Table S1 for details on data sources.

Table S29: Mortality rates by sex, age and education - Suicides

| Country | Education | 20-29 | 30-39 | 40-49 | 50-59 | 60-69 | 70-79 | 80+ |
|---|---|---|---|---|---|---|---|---|
| | | | | Women | | | | |
| Argentina 2011 | Low | 0.05 | 0.04 | 0.04 | 0.03 | 0.03 | 0.02 | 0.03 |
| | High | 0.03 | 0.03 | 0.03 | 0.04 | 0.02 | 0.02 | 0.02 |
| Brazil 2011 | Low | 0.03 | 0.03 | 0.03 | 0.03 | 0.02 | 0.03 | 0.02 |
| | High | 0.02 | 0.03 | 0.03 | 0.04 | 0.03 | 0.02 | 0.01 |
| Chile 2018 | Low | 0.13 | 0.06 | 0.04 | 0.05 | 0.02 | 0.02 | 0.00 |
| | High | 0.05 | 0.05 | 0.05 | 0.04 | 0.03 | 0.02 | 0.04 |
| Colombia 2019 | Low | 0.05 | 0.03 | 0.02 | 0.02 | 0.02 | 0.02 | 0.00 |
| | High | 0.04 | 0.02 | 0.02 | 0.02 | 0.02 | 0.01 | 0.00 |
| Ecuador 2011 | Low | 0.07 | 0.03 | 0.04 | 0.03 | 0.02 | 0.02 | 0.03 |
| | High | 0.05 | 0.02 | 0.03 | 0.02 | 0.03 | 0.00 | 0.00 |
| Mexico 2011 | Low | 0.02 | 0.01 | 0.01 | 0.01 | 0.01 | 0.00 | 0.00 |
| | High | 0.01 | 0.01 | 0.01 | 0.01 | 0.01 | 0.00 | 0.00 |
| Peru 2018 | Low | 0.02 | 0.00 | 0.01 | 0.00 | 0.00 | 0.00 | 0.00 |
| | High | 0.00 | 0.00 | 0.00 | 0.00 | 0.00 | 0.00 | 0.00 |
| | | | | Men | | | | |
| Argentina 2011 | Low | 0.29 | 0.17 | 0.12 | 0.14 | 0.18 | 0.24 | 0.33 |
| | High | 0.15 | 0.09 | 0.08 | 0.10 | 0.12 | 0.15 | 0.23 |
| Brazil 2011 | Low | 0.15 | 0.14 | 0.13 | 0.12 | 0.12 | 0.15 | 0.16 |
| | High | 0.09 | 0.09 | 0.10 | 0.11 | 0.13 | 0.12 | 0.20 |
| Chile 2018 | Low | 0.45 | 0.45 | 0.34 | 0.30 | 0.30 | 0.26 | 0.34 |
| | High | 0.18 | 0.21 | 0.20 | 0.19 | 0.16 | 0.23 | 0.22 |
| Colombia 2019 | Low | 0.23 | 0.16 | 0.14 | 0.15 | 0.16 | 0.13 | 0.15 |
| | High | 0.13 | 0.10 | 0.12 | 0.12 | 0.14 | 0.14 | 0.06 |
| Ecuador 2011 | Low | 0.25 | 0.14 | 0.12 | 0.16 | 0.18 | 0.15 | 0.13 |
| | High | 0.15 | 0.08 | 0.06 | 0.07 | 0.05 | 0.12 | 0.12 |
| Mexico 2011 | Low | 0.11 | 0.07 | 0.07 | 0.05 | 0.05 | 0.06 | 0.07 |
| | High | 0.03 | 0.03 | 0.03 | 0.03 | 0.05 | 0.05 | 0.06 |
| Peru 2018 | Low | 0.02 | 0.01 | 0.01 | 0.01 | 0.01 | 0.00 | 0.01 |
| | High | 0.01 | 0.01 | 0.00 | 0.00 | 0.00 | 0.00 | 0.00 |

*Note:* Authors' calculations based on census and death certificate data. See Appendix Table S1 for details on data sources.

Table S30: Mortality rates ratios by sex, age and education - Suicides

| Country | 20-29 | 30-39 | 40-49 | 50-59 | 60-69 | 70-79 | 80+ |
|---|---|---|---|---|---|---|---|
| | | | Women | | | | |
| Argentina 2011 | 1.72 | 1.60 | 1.38 | 0.78 | 1.28 | 1.09 | 1.69 |
| Brazil 2011 | 1.40 | 1.09 | 0.90 | 0.80 | 0.85 | 1.09 | 1.36 |
| Chile 2018 | 2.85 | 1.14 | 0.81 | 1.02 | 0.63 | 0.63 | 0.13 |
| Colombia 2019 | 1.27 | 1.49 | 1.22 | 0.90 | 0.70 | 1.83 | - |
| Ecuador 2011 | 1.50 | 2.00 | 1.56 | 1.55 | 0.72 | - | - |
| Mexico 2011 | 1.41 | 1.30 | 1.18 | 0.74 | 0.70 | 1.04 | - |
| Peru 2018 | - | - | 6.91 | - | - | - | - |
| Median | 1.46 | 1.40 | 1.22 | 0.85 | 0.71 | 1.09 | 1.36 |
| Max/Min | 2.24 | 1.83 | 8.54 | 2.10 | 2.04 | 2.89 | 13.04 |
| Men | | | | | | | |
| Argentina 2011 | 1.92 | 1.81 | 1.44 | 1.49 | 1.46 | 1.63 | 1.42 |
| Brazil 2011 | 1.67 | 1.47 | 1.27 | 1.11 | 0.93 | 1.22 | 0.81 |
| Chile 2018 | 2.56 | 2.16 | 1.70 | 1.61 | 1.92 | 1.15 | 1.55 |
| Colombia 2019 | 1.83 | 1.52 | 1.15 | 1.20 | 1.17 | 0.98 | 2.67 |
| Ecuador 2011 | 1.70 | 1.73 | 1.99 | 2.16 | 3.29 | 1.20 | 1.10 |
| Mexico 2011 | 3.24 | 2.11 | 2.39 | 1.56 | 1.12 | 1.13 | 1.13 |
| Peru 2018 | 4.03 | 1.15 | 3.86 | 5.15 | 3.21 | - | - |
| Median | 1.92 | 1.73 | 1.70 | 1.56 | 1.46 | 1.17 | 1.27 |
| Max/Min | 2.41 | 1.88 | 3.36 | 4.62 | 3.53 | 1.67 | 3.30 |

*Note:* Authors' calculations based on census and death certificate data. See Appendix Table S1 for details on data sources.

#### Homicides

Figure S8: Homicides

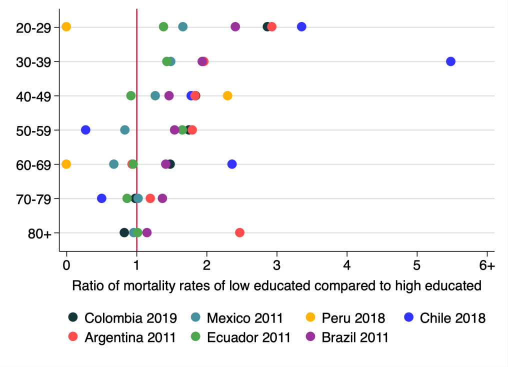

Women

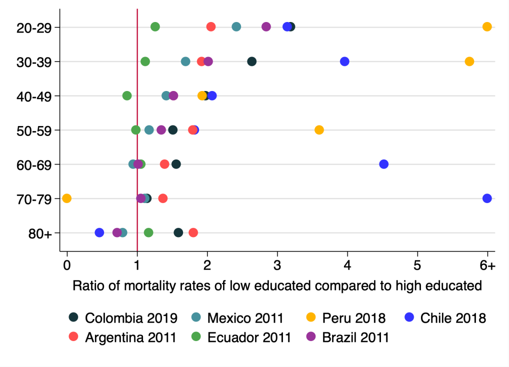

Men

*Note:* Authors' calculations based on census and death certificate data. See Appendix Table S1 for details on data sources.

Table S31: Mortality rates by sex, age and education - Homicides

| Country | Education | 20-29 | 30-39 | 40-49 | 50-59 | 60-69 | 70-79 | 80+ |
|---|---|---|---|---|---|---|---|---|
| | | | | Women | | | | |
| Argentina 2011 | Low | 0.03 | 0.02 | 0.02 | 0.02 | 0.01 | 0.02 | 0.02 |
| | High | 0.01 | 0.01 | 0.01 | 0.01 | 0.01 | 0.02 | 0.01 |
| Brazil 2011 | Low | 0.13 | 0.10 | 0.05 | 0.03 | 0.03 | 0.03 | 0.03 |
| | High | 0.05 | 0.05 | 0.04 | 0.02 | 0.02 | 0.02 | 0.03 |
| Chile 2018 | Low | 0.03 | 0.03 | 0.01 | 0.00 | 0.01 | 0.00 | 0.01 |
| | High | 0.01 | 0.01 | 0.01 | 0.01 | 0.01 | 0.01 | 0.00 |
| Colombia 2019 | Low | 0.17 | 0.12 | 0.08 | 0.04 | 0.03 | 0.03 | 0.03 |
| | High | 0.06 | 0.06 | 0.04 | 0.03 | 0.02 | 0.03 | 0.03 |
| Ecuador 2011 | Low | 0.07 | 0.06 | 0.03 | 0.05 | 0.04 | 0.04 | 0.12 |
| | High | 0.05 | 0.04 | 0.03 | 0.03 | 0.04 | 0.04 | 0.12 |
| Mexico 2011 | Low | 0.05 | 0.04 | 0.03 | 0.02 | 0.02 | 0.02 | 0.03 |
| | High | 0.03 | 0.02 | 0.02 | 0.03 | 0.03 | 0.02 | 0.03 |
| Peru 2018 | Low | 0.00 | 0.00 | 0.00 | 0.00 | 0.00 | 0.01 | 0.00 |
| | High | 0.00 | 0.00 | 0.00 | 0.00 | 0.00 | 0.00 | 0.00 |
| | | | | Men | | | | |
| Argentina 2011 | Low | 0.20 | 0.15 | 0.09 | 0.06 | 0.06 | 0.08 | 0.12 |
| | High | 0.10 | 0.08 | 0.06 | 0.04 | 0.04 | 0.06 | 0.07 |
| Brazil 2011 | Low | 1.74 | 0.96 | 0.53 | 0.32 | 0.22 | 0.16 | 0.14 |
| | High | 0.61 | 0.48 | 0.35 | 0.23 | 0.22 | 0.15 | 0.20 |
| Chile 2018 | Low | 0.23 | 0.24 | 0.09 | 0.05 | 0.04 | 0.05 | 0.01 |
| | High | 0.07 | 0.06 | 0.04 | 0.03 | 0.01 | 0.01 | 0.02 |
| Colombia 2019 | Low | 2.28 | 1.57 | 0.83 | 0.45 | 0.29 | 0.16 | 0.12 |
| | High | 0.71 | 0.59 | 0.42 | 0.30 | 0.18 | 0.14 | 0.08 |
| Ecuador 2011 | Low | 0.63 | 0.56 | 0.37 | 0.27 | 0.16 | 0.19 | 0.28 |
| | High | 0.49 | 0.50 | 0.43 | 0.27 | 0.15 | 0.00 | 0.24 |
| Mexico 2011 | Low | 0.57 | 0.49 | 0.33 | 0.20 | 0.14 | 0.10 | 0.12 |
| | High | 0.24 | 0.29 | 0.23 | 0.17 | 0.15 | 0.09 | 0.15 |
| Peru 2018 | Low | 0.05 | 0.02 | 0.01 | 0.02 | 0.01 | 0.00 | 0.00 |
| | High | 0.01 | 0.00 | 0.00 | 0.01 | 0.00 | 0.00 | 0.00 |

*Note:* Authors' calculations based on census and death certificate data. See Appendix Table S1 for details on data sources.

Table S32: Mortality rates ratios by sex, age and education - Homicides

| Country | 20-29 | 30-39 | 40-49 | 50-59 | 60-69 | 70-79 | 80+ |
|---|---|---|---|---|---|---|---|
| Women | | | | | | | |
| Argentina 2011 | 2.93 | 1.96 | 1.84 | 1.79 | 0.94 | 1.20 | 2.48 |
| Brazil 2011 | 2.41 | 1.94 | 1.46 | 1.55 | 1.42 | 1.37 | 1.15 |
| Chile 2018 | 3.36 | 5.49 | 1.78 | 0.27 | 2.36 | 0.51 | - |
| Colombia 2019 | 2.87 | 1.94 | 1.85 | 1.75 | 1.48 | 0.99 | 0.83 |
| Ecuador 2011 | 1.38 | 1.44 | 0.92 | 1.65 | 0.95 | 0.87 | 1.02 |
| Mexico 2011 | 1.66 | 1.49 | 1.27 | 0.83 | 0.67 | 1.02 | 0.96 |
| Peru 2018 | - | - | 2.30 | - | - | - | - |
| Median | 2.64 | 1.94 | 1.78 | 1.60 | 1.19 | 1.00 | 1.02 |
| Max/Min | 2.43 | 3.82 | 2.50 | 6.59 | 3.50 | 2.71 | 3.00 |
| Men | | | | | | | |
| Argentina 2011 | 2.05 | 1.92 | 1.52 | 1.79 | 1.39 | 1.37 | 1.80 |
| Brazil 2011 | 2.84 | 2.01 | 1.52 | 1.35 | 1.02 | 1.05 | 0.72 |
| Chile 2018 | 3.15 | 3.96 | 2.07 | 1.82 | 4.52 | 7.24 | 0.47 |
| Colombia 2019 | 3.19 | 2.64 | 1.98 | 1.51 | 1.56 | 1.14 | 1.59 |
| Ecuador 2011 | 1.26 | 1.12 | 0.86 | 0.98 | 1.05 | - | 1.17 |
| Mexico 2011 | 2.42 | 1.70 | 1.42 | 1.17 | 0.95 | 1.11 | 0.80 |
| Peru 2018 | 9.51 | 5.75 | 1.93 | 3.60 | - | - | - |
| Median | 2.84 | 2.01 | 1.52 | 1.51 | 1.22 | 1.14 | 0.98 |
| Max/Min | 7.53 | 5.13 | 2.41 | 3.67 | 4.77 | 6.86 | 3.87 |

*Note:* Authors' calculations based on census and death certificate data. See Appendix Table S1 for details on data sources.
